# A Survey of Copy Number Variants Associated with Neurodevelopmental Disorders in a Large-Scale, Multi-Ancestry Biobank

**DOI:** 10.1101/2021.06.09.21258554

**Authors:** Rebecca Birnbaum, Behrang Mahjani, Ruth J.F. Loos, Andrew J. Sharp

**Affiliations:** Department of Psychiatry, Icahn School of Medicine at Mount Sinai; Department of Genetics and Genomic Sciences, Icahn School of Medicine at Mount Sinai; Seaver Autism Center for Research and Treatment, Icahn School of Medicine at Mount Sinai; Charles Bronfman Institute for Personalized Medicine, Icahn School of Medicine at Mount Sinai; Novo Nordisk Foundation Center for Basic Metabolic Research, University of Copenhagen, Denmark; The Mindich Child Health and Development Institute, Icahn School of Medicine at Mount Sinai

**Keywords:** Copy Number Variant, Biobank, Neurodevelopmental Disorders, Autism Spectrum Disorders, Schizophrenia

## Abstract

**BACKGROUND:** Past clinical genetic studies have identified rare, copy number variants (CNVs) as risk factors for multiple neurodevelopmental disorders (NDD), including autism spectrum disorder and schizophrenia. However, the broad, clinical characterization of these NDD-CNVs in large population cohorts, especially of diverse ancestry, is relatively understudied. We characterized the clinical presentation of NDD-CNVs in the Bio*Me* biobank, comprising ∼25,000 individuals across diverse ancestry, medical and neuropsychiatric clinical presentation, with a mean age of 50.3 years.

**METHODS:** Individuals within the Bio*Me* biobank harboring NDD-CNVs were identified using a consensus of two CNV calling algorithms, based on whole-exome sequencing and genotype array data, followed by a series of novel, *in-silico* clinical assessments.

**RESULTS:** The overall prevalence of a set of 64 NDD-CNVs was calculated at ∼2.5%, with prevalence varying by locus, corroborating the presence of some relatively, highly-prevalent NDD-CNVs (i.e., 15q11.2 deletion/duplication, 2q13(*NPHP1*) deletion/duplication). An aggregate set of rare, NDD-CNVs were enriched for congenital disorders (OR=1.8, p-value=0.02) and major depressive disorders (OR=1.3, p-value=0.04) in multi-ancestry analyses, and major depressive-disorder in an African ancestry-stratified group (OR=1.8, p-value=0.01). In a meta-analysis of medical diagnoses (n=195 hierarchically-clustered diagnostic codes), an aggregated set of rare, NDD-CNVs was significantly associated with obstructive sleep apnea (Z-score=3.6 p=3.2×10^−4^). Further, an aggregated set of rare, NDD-CNVs was associated with increased body mass index (BMI) in a multi-ancestry analysis (Beta=0.14, p-value=0,04), and in Hispanic-stratified analyses (Beta=0.30, p-value=4.2×10^−3^). For 38 common serum laboratory tests, there was no identified association with an aggregate set of NDD-CNVs.

**CONCLUSION:** The current analyses elucidated clinical features of individuals harboring NDD-CNVs, in a large-scale, multi-ancestry biobank, identifying enrichments for congenital disorders and major depressive disorder, as well as identifying associations with obesity-related phenotypes, obstructive sleep apnea and increased BMI. Future recall of individuals harboring NDD-CNVs will allow for further clinical assessments beyond the electronic health records (EHR) presently analyzed, including neurocognitive and neuroimaging outcomes.

## INTRODUCTION

Clinical genomic investigations to date have identified rare copy number variants (CNVs, i.e. genomic microdeletions or microduplications >1kb) considered pathogenic for neurodevelopmental disorders (NDD), including schizophrenia (SCZ), autism spectrum disorder (ASD), and intellectual disability (ID).^1-3^ Initial genomic studies of CNVs in NDD utilized chromosomal microarray analyses, while most recent discovery and validation efforts have leveraged genome-wide association analyses (GWAS), with CNVs called from genotyping arrays.^2-6^ Among NDD-pathogenic CNVs, eight CNVs are reported to occur at significantly increased frequencies in schizophrenia compared to controls, including the 22q11.2 deletion underlying velocardiofacial syndrome, which is among the most well-studied and clinically characterized.^2, 7^ Nearly all NDD-CNVs have a greater effect on disease risk compared to common genetic variants. For example, the reported odds ratios (OR) of schizophrenia-associated-CNVs range from 3-60, whereas the OR conferred by most GWAS-significant single nucleotide polymorphisms (SNP) is less than 1.2.^2, 8^ NDD-pathogenic CNVs have been characterized in clinical studies and population cohorts, to quantify clinical features of developmental delay, physical stigmata and neuropsychiatric effects, and have been established as both inherited and *de novo* risk factors. ^9-12^

Despite considerable investigation of NDD-pathogenic CNVs to date, significant gaps in their clinical characterization remain. First, pleiotropy, or non-specificity of NDD-pathogenic CNVs remains unexplained, that is the same CNV may confer risk for *multiple* neurodevelopmental disorders.^13^ Second, the variable penetrance and expressivity remain poorly understood, that is among NDD-CNV carriers, developmental or neuropsychiatric symptomatology ranges from unaffected to severely affected.^14^ Some studies suggest a role of ‘background’ genetic risk, polygenic risk derived from common risk alleles, in interacting with NDD-pathogenic CNVs to influence penetrance; in other reports, there is evidence of a ‘two-hit’ model whereby a large NDD-CNV event exacerbated neurodevelopmental phenotypes, in association with other large deletions or duplications.^15-18^ Most clinical reports of NDD-CNVs to date have focused on neuropsychiatric traits, absent overall medical co-morbidities. Lastly, most population cohorts/registries, or large-scale biobank studies to date of CNVs have been limited to European ancestry, limiting overall clinical generalizability.^19-22^

The precise pathogenic factor within many NDD-pathogenic CNV regions remains unknown, whether gene dosage effect of protein-coding gene(s), non-coding species (i.e., microRNA) or other modifying, regulatory factors.^23, 24^ Further investigation of CNV carriers may elucidate not only underlying biological factors or mechanisms of pathogenesis, but also accelerate potential translation efforts and more precise therapeutic strategies.

Clinical investigation of CNV carriers is challenging however, given their rare frequency and requisite access to large-scale clinical and genetic resources, concurrently. Several research consortia focus on individual CNVs (i.e., 22q11 deletion, 3q29 deletion or 16p11.2), while other consortia focus on CNVs in aggregate, for example the Deciphering Developmental Disorders project or Enhancing Imaging Genetics through Meta-Analysis (ENIGMA-CNV), the latter investigating neuroimaging outcomes.^25-27^ The study of multiple NDD-pathogenic CNVs is advantageous since it permits inter-CNV comparison and assessment of composite clinical outcomes.

The current study leveraged a robust clinical genetics resource, the “Bio*Me*” biobank, a genetics repository linked to the electronic health records (EHR) of approximately 25,000 individuals recruited across medical specialties and across ancestry and socio-economic background.^28, 29^ Notably, the Bio*Me* biobank is multi-ancestry, enabling diverse query across ancestry (in contrast to most biobanks studies to date, which have focused primarily on European ancestry). In a *forward* genetics approach, individuals harboring NDD-pathogenic CNVs were identified without phenotypic ascertainment bias, followed by a series of novel *in-silico* clinical assessments. The genetics repository of the biobank enabled the identification of individuals whom harbor NDD-CNVs, while the EHR data enabled investigation of clinical indices, including diagnostic codes, laboratory serum tests, and clinical descriptive data contained within documented encounters.

## METHODS

### “BioMe” Biobank

Participants were recruited throughout the Mount Sinai healthcare system, as per a protocol approved by the local Institutional Review Board (IRB), initiated in 2007. Participants were recruited across age and ancestry, and from clinics throughout the healthcare system of diverse medical and neuropsychiatric specialty (See Supplementary Table 1, Supplementary Methods). In providing informed consent, Biobank participants authorized access to their de-identified healthcare records and also donated a blood sample for extraction of genetic material for research purposes. As per the approved protocol, no disclosure/feedback of genetic results would be provided, as the analyses were for research purposes. Participants were offered the option to consent to be re-contacted for future research studies.

### Sample Genotyping and Exome Sequencing

As previously described, Bio*Me* biobank participants were genotyped on the Illumina Global Screening Array (GSA-24v1.0) and exome sequencing was performed at the Regeneron Genetics Center.^29^ (See Supplementary Methods for details of genotyping and exome sequencing QC and filtering).

### CNV Calling

The 64 CNVs called in the current analysis, herein termed “NDD-CNVs”, were reported in previous biobank reports (i.e., UK Biobank and Geisinger DiscoverEHR), described as set of CNVs ‘pathogenic’ for neurodevelopment disorders (NDD), culled from 92 CNVs in 47 genomic locations, including reciprocal deletions/duplications.^5, 6, 19, 20^ ^30, 31^ A CNV was called based on the union, or consensus of two CNV calling algorithms (each using a different type of sequence data), a whole exome-sequenced based method, Lattice-Aligned Mixture Models (CLAMMS), and a genotype array-based method, PennCNV. (See Supplementary Methods for details of each CNV calling method, QC and filtering).^32, 33^ Only Bio*Me* samples with *both* WES-based CLAMMS calls and array-based PennCNV calls, subsequent to QC (n=24,877 samples) were included in NDD-CNV analyses and downstream phenotypic enrichment and association analyses (See Supplementary Methods). As per previous biobank reports, individuals were designated as positive for a CNV, if breakpoints overlapped at least 50% of the defined critical region, and for single gene CNV deletions (i.e., *NRXN1* deletions) intersected at least one exon.^19, 20^

### Enrichment for Neuropsychiatric Disorders

For enrichment analyses, samples without phenotype data were excluded, as well as samples with less than two clinical encounters, to increase the reliability of phenotypic analyses, yielding n=22,279 individuals for the analyses of enrichment of neuropsychiatric disorders. A Fisher’s exact test was applied to test for statistical significance of enrichment of an aggregate, set of rare, NDD-CNVs (excluding the common 15q11.2 del/duplication) for neurodevelopmental and neuropsychiatric disease categories.

### Phenome-Wide Association Analyses (“PheWAS”)

To reduce potential confounding effects of relatedness, a random individual from each related pair with more than second^-^degree relatedness (kinship coefficient>0.0885) was excluded, as estimated based on genotype data (See Supplementary Methods).

PheWAS was conducted using ‘REGENIE’, a machine-learning method, especially developed for rare variant analysis of binary (case-control) traits with unbalanced case-control ratios.^34^ International Classification of Diseases codes (ICD9 and ICD10) were mapped to hierarchically-clustered ‘phecodes’ and for each phecode, the number of cases and controls determined, longitudinally, with a case defined as at least 2 counts, a control as 0, while a count of 1 were set to missing for the phecode.^35, 36^ Phecodes were filtered for a minimum of 100 cases for each ancestry-stratified group, European, African, and Hispanic, to ensure robust analyses, so that only the more prevalent n=195 phecodes, of 1,736 phecodes across the biobank cohort, were tested in the current NDD-CNV PheWAS (See Supplementary Methods). Using REGENIE, each phecode was regressed onto CNV (binary) status with covariates of: age, sex, ancestry, principal components (PCs) 1-5 of the genotype data, and density of electronic health records (See Supplementary Methods). For each PheWAS conducted using REGENIE, Bonferroni-correction was applied to control for multiple testing. In addition to ancestry-stratified analyses, multi-ancestry meta-analysis, was conducted using METAL^37^

### Association Analyses with Quantitative Outcome

Median BMI and serum lab values were inverse normal transformed, and regressed onto NDD-CNV status (binary variable), adjusting for covariates of age, sex, ancestry, PCs 1-5 of the genotype data, and density of electronic health records (See Supplementary Methods).

## RESULTS

The Bio*Me* biobank cohort utilized in the current CNV study (n=24,877 individuals) is multi-ancestry (31.7% European, 24% African, 34.3% Hispanic, 10% Other), consisting of a range of ages at enrollment, but skewed towards older adults (mean age at enrollment: 50.3 years), with few pediatric participants (2.5%<18 years), and more women (59%) than men (Table 1). To initiate the current investigation, individuals harboring NDD-CNVs (any of 64 NDD-CNVs) were identified within the “Bio*Me*” Biobank.

**Table 1:**
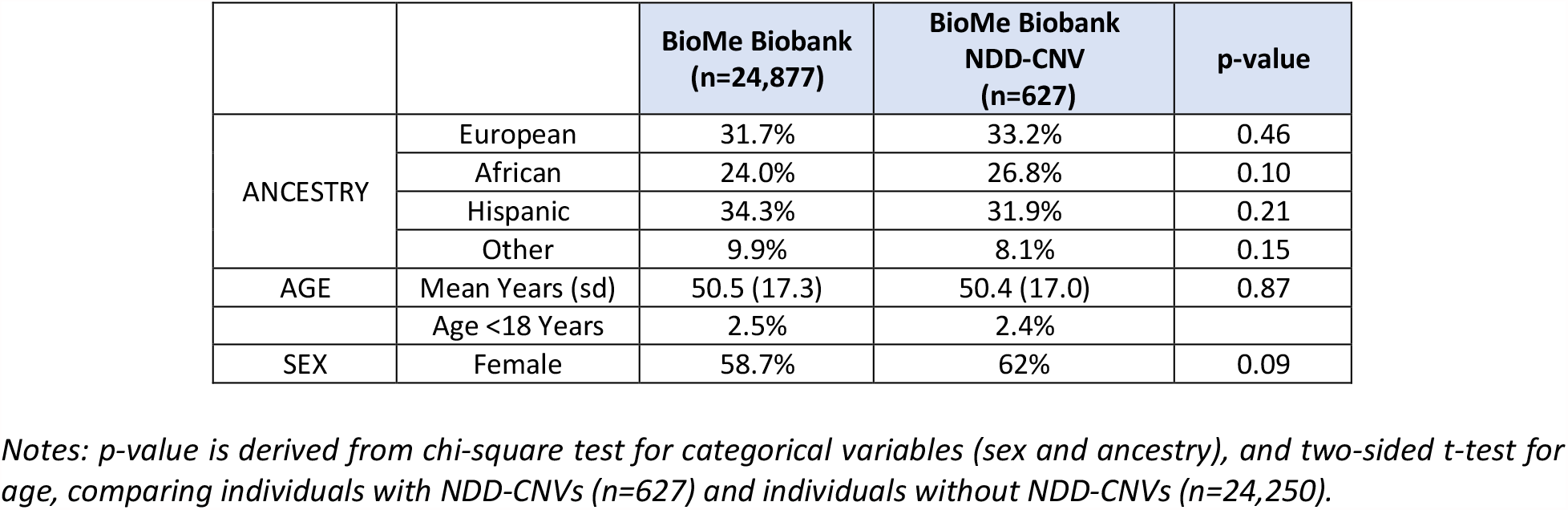
BioMe Biobank Demographic Overview: Demographics of biobank participants included in current analysis (n=24,877) and subset harboring NDD-CNVs (n=627)

### CNV Prevalence

CNVs were called from the consensus of two CNV calling algorithms, a whole exome sequencing-based method (CLAMMS), and a genotype array-based method, PennCNV, yielding the result that 2.5% of individuals (n=627 individuals) within the biobank harbor a NDD-CNV (Table 2).^32, 33^

**TABLE 2:** NDD-CNVs in the “Bio*Me*” Biobank: Each of 64 NDD-CNVs genotyped in the Biobank using a consensus of two CNV calling algorithms

|  | LOCATION (hg19) | GENES (n) | BioMe (n) | PREVALENCE (%) |
| --- | --- | --- | --- | --- |
| TAR del | chr1:145,39–145,81 | 17 | 5 | 0.020 |
| TAR dup | chr1:145,39–145,81 | 17 | 18 | 0.072 |
| 1q21.1del | chr1:146,53–147,39 | 9 | 8 | 0.032 |
| 1q21.1dup | chr1:146,53–147,39 | 9 | 3 | 0.012 |
| NRXN1 del | chr2:50,14–51,26 | 1 | 3 | 0.012 |
| 2q11.2del | chr2:96,74–97,68 | 22 | 2 | 0.008 |
| 2q11.2dup | chr2:96,74–97,68 | 22 | 1 | 0.004 |
| 2q13del(NPHP1) | chr2:110,86–110,98 | 3 | 81 | 0.326 |
| 2q13dup(NPHP1) | chr2:110,86–110,98 | 3 | 62 | 0.249 |
| 2q13del | chr2:111,39–112,01 | 3 | 3 | 0.012 |
| 2q13dup | chr2:111,39–112,01 | 3 | 8 | 0.032 |
| 2q21.1del | chr2:131,48–131,93 | 5 | 9 | 0.036 |
| 2q21.1dup | chr2:131,48–131,93 | 5 | 5 | 0.020 |
| 3q29del | chr3:195,72–197,35 | 28 | 1 | 0.004 |
| 3q29dup | chr3:195,72–197,35 | 28 | 0 | 0 |
| Sotos 5q35del | chr5:175,72–177,05 | 39 | 0 | 0 |
| 5q35dup | chr5:175,72–177,05 | 39 | 0 | 0 |
| 7q11.23 del | chr7:72,74–74,14 | 26 | 1 | 0.004 |
| WBS 7q11.23 dup | chr7:72,74–74,14 | 26 | 0 | 0 |
| 7q11.23dup distal | chr7:75,14–76,06 | 16 | 0 | 0 |
| 8p23.1del | chr8:8,10–11,87 | 35 | 0 | 0 |
| 8p23.1dup | chr8:8,10–11,87 | 35 | 0 | 0 |
| 10q11.21q11.23del | chr10:49,39–51,06 | 19 | 1 | 0.004 |
| 10q11.21q11.23dup | chr10:49,39–51,06 | 19 | 2 | 0.008 |
| 10q23del | chr10:82,05–88,93 | 29 | 0 | 0 |
| 10q23dup | chr10:82,05–88,93 | 29 | 1 | 0.004 |
| 13q12del(CRYL1) | chr13:20,98–21,10 | 2 | 8 | 0.032 |
| 13q12dup(CRYL1) | chr13:20,98–21,10 | 2 | 0 | 0 |
| 13q12.12del | chr13:23,56–24,88 | 10 | 4 | 0.016 |
| 13q12.12dup | chr13:23,56–24,88 | 10 | 9 | 0.036 |
| 15q11.2del | chr15:22,81–23,09 | 5 | 60 | 0.241 |
| 15q11.2dup | chr15:22,81–23,09 | 5 | 163 | 0.655 |
| PW/AS 15q11.2q13.1 BP1-3 del | chr15:23,68–28,39 | 116 | 0 | 0 |
| PW/AS 15q11.2q13.1 BP1-3 dup | chr15:23,68–28,39 | 116 | 0 | 0 |
| 15q11q13del BP3- | chr15:29,16–30,38 | 4 | 1 | 0.004 |
| 15q11q13dup BP3- | chr15:29,16–30,38 | 4 | 3 | 0.012 |
| 15q11q13dup BP3-BP5 | chr15:29,16–32,46 | 17 | 0 | 0 |
| 15q13.3del | chr15:31,08–32,46 | 8 | 5 | 0.020 |
| 15q13.3dup | chr15:31,08–32,46 | 8 | 5 | 0.020 |
| 15q13.3del(CHRNA7) | chr15:32,02–32,46 | 1 | 5 | 0.020 |
| 15q13.3dup(CHRNA7) | chr15:32,02–32,46 | 1 | 42 | 0.169 |
| 15q24del | chr15:72,90–78,15 | 77 | 0 | 0 |
| 15q24dup | chr15:72,90–78,15 | 77 | 0 | 0 |
| 16p13.11del | chr16:15,51–16,29 | 7 | 13 | 0.052 |
| 16p13.11dup | chr16:15,51–16,29 | 7 | 39 | 0.157 |
| 16p12.1del | chr16:21,95–22,43 | 8 | 2 | 0.008 |
| 16p12.1dup | chr16:21,95–22,43 | 8 | 14 | 0.056 |
| 16p11.2distal del | chr16:28,82–29,05 | 11 | 5 | 0.020 |
| 16p11.2distal dup | chr16:28,82–29,05 | 11 | 4 | 0.016 |
| 16p11.2del | chr16:29,65–30,20 | 30 | 15 | 0.060 |
| 16p11.2dup | chr16:29,65–30,20 | 30 | 4 | 0.016 |
| 17p12del | chr17:14,14–15,43 | 8 | 6 | 0.024 |
| 17p12dup | chr17:14,14–15,43 | 8 | 10 | 0.040 |
| Smith Magenis | chr17:16,81–20,21 | 59 | 0 | 0 |
| Potocki-Lupski | chr17:16,81–20,21 | 59 | 0 | 0 |
| 17q11.2del(NF1) | chr17:29,12–30,27 | 19 | 0 | 0 |
| 17q11.2dup(NF1) | chr17:29,12–30,27 | 19 | 0 | 0 |
| 17q12del | chr17:34,81–36,22 | 17 | 4 | 0.016 |
| 17q12dup | chr17:34,81–36,22 | 17 | 4 | 0.016 |
| 17q21.31del | chr17:43,70–44,29 | 10 | 0 | 0 |
| 22q11.2del | chr22:19,04–21,47 | 61 | 1 | 0.004 |
| 22q11.2dup | chr22:19,04–21,47 | 61 | 7 | 0.028 |
| 22q11.2distal del | chr22:21,92–23,65 | 26 | 0 | 0 |
| 22q11.2distal dup | chr22:21,92–23,65 | 26 | 0 | 0 |

Prevalence varied by CNV locus and type, notable for six highly-prevalent CNVs: chr15q11.2 deletion and duplication, chr2q13(*NPHP1*) deletion and duplication, chr15q13.3 (*CHRNA7*) duplication, and chr16p13.11 duplication (Supplementary Table 2). Only one carrier of the 22q11.2 hemizygous deletion was identified, and for 21 CNVs, there were no carriers identified (i.e., chr15-PWS-DUP, chr17 Potocki-Lupski syndrome, 17q11.2(*NF1*)DEL). The prevalence for each NDD-CNV in Bio*Me* is compared with previously-reported UK Biobank and DiscoverEHR CNV analyses (Supplementary Table 2). ^19, 20^

Overall, within the biobank cohort, the demographic variables of age, sex and ancestry, did not differ significantly for the subset of NDD-CNV carriers, compared to individuals without NDD-CNVs (Table 1). In addition to the multi-ancestry composition, a notable demographic feature of the NDD-CNV carriers is the mean of 50 years of age, similar to the overall Bio*Me* cohort. Therefore, in the current analysis, NDD-CNVs were not surveyed in a childhood or developmental context, specifically, but rather mostly in older adults. In assessing relatedness among NDD-CNV carriers, 99 pairs of relatives of at least second-degree were identified: 17 pairs in whom both relatives harbored NDD-CNVs, and 82 pairs in which one, but not both relatives, harbored a NDD-CNV (Supplementary Table 3). Furthermore, among NDD-CNV carriers within the biobank, n=20 individuals harbored two NDD-CNVs, each.

### Enrichment of Neuropsychiatric Disorders

Next, individuals harboring rare, NDD-CNVs were tested for enrichment of neurodevelopmental disorders (Table 3, Supplementary Table 4 and Supplementary Methods). The aggregate set of NDD-CNV carriers (n=376) were enriched for congenital disorders (Fisher’s exact two-sided p-value=0.02, OR=1.8), but not schizophrenia (p=1) nor seizure disorder (p=1). In ancestry-stratified analyses of congenital disorders, NDD-CNVs were not enriched among the major three ancestry subgroups (European p=1, African p=0.16, Hispanic p=0.13) but significant for the ‘other’ ancestry subgroup (p-value=0.005, OR=6.9) but with a wide-ranging 95% confidence interval, owing to small group size (n=54 combined Asian, Native American and other).

**TABLE 3:** Enrichment for Neurodevelopmental/Neuropsychiatric Disorders: Among carriers of NDD-CNVs, enrichment for neurodevelopmental and neuropsychiatric disorders

| **(NEURO)DEVELOPMENTAL DISORDER** | **n** | **p-value** | **OR** | **95% CI** |
| --- | --- | --- | --- | --- |
| SCZ/OTHER SCZ PSYCHOSIS | 412 | 1 | 0.9 | 0.3-1.9 |
| ASD/ID | 29 | 0.09 | 4.3 | 0.5-17.3 |
| CONGENITAL DISORDERS | 639 | 0.02 | 1.8 | 1.1-2.9 |
| EPILEPSY/SEIZURE | 342 | 1 | 0.9 | 0.3-2.1 |

| **ADULT-ONSET NEUROPSYCHIATRIC DISORDER** | **n** | **p-value** | **OR** | **95% CI** |
| --- | --- | --- | --- | --- |
| BIPOLAR DISORDER | 329 | 0.83 | 1.08 | 0.4-2.4 |
| MAJOR DEPRESSIVE DISORDER | 3668 | 0.04 | 1.31 | 1-1.7 |

*Notes:*
-SCZ: schizophrenia; ASD: Autism Spectrum Disorders; ID: Intellectual Disability
-See Supplementary Table 4 for description of each disorder, by ICD code, and for tabulation of NDD-CNVs tested.
-Fisher's exact test for NDD CNV enrichment, by disorder, reported two-sided p-value, OR, 95% CI indicated.

Enrichment for ASD/ID trended towards significance (p=0.09) but with notably few cases represented in Bio*Me;* within the Bio*Me* cohort, the prevalence of childhood developmental disorders is markedly low (Bio*Me* ASD/ID prevalence=0.13%) owing to few pediatric participants, compared to adult-onset disorders, which better approximate overall population prevalence (i.e. Bio*Me* SCZ prevalence=1.1%).

Enrichment for adult-onset mood disorders was also queried, and NDD-CNVs were found to be enriched for major depressive disorder (MDD) (Fisher’s exact two-sided p-value=0.04, OR=1.3) with the enrichment for MDD driven by individuals of African ancestry (two-sided p-value=0.01, OR=1.8). In contrast, there was no NDD-CNV enrichment for bipolar disorder (two-sided p-value=0.83).

### Association with ICD-Diagnosis Codes (‘PheWAS’)

To investigate phenotypic associations in the biobank broadly, beyond neuropsychiatric disorders, an agnostic phenome-wide association (“PheWAS”) was next applied, to test NDD-CNV association with ICD codes, coded medical diagnoses across organ systems.^38, 39^ Here, PheWAS tested 195 hierarchically-clustered ICD codes (i.e. ‘phecodes’’) for association with each of five NDD-CNV sets, (1) an aggregated set of rare, NDD-CNVs (n=225, excluding 15q11.2 deletion/duplication and 2q13(*NPHP1*) deletion/duplication), and prevalent, individual NDD-CNVs: (2) 15q13.3 duplication (*CHRNA7*) (n=36); (3) 16p13.11 duplication (n=34); (4) 2q13(*NPHP1*) deletion (n= 63); (5) 2q13(*NPHP1*) duplication (n=53). Each NDD-CNV PheWAS was stratified by ancestry (European, African, Hispanic), as well as combined in a multi-ancestry meta-analysis.

Overall, PheWAS across the NDD-CNV sets, yielded several significant associations (Figure 1). The aggregated set of rare, NDD-CNVs were significantly associated with ‘Obstructive sleep apnea’ (Z-score=3.6, p=3.24×10-4) and ‘Urinary tract infection’ (Z-score=3.5, p=4.4×10-4), while subthreshold association was noted with ‘essential hypertension’ (Z-score=-3.00, p=0.003) and ‘acute renal failure (Z-score=2.8, p=0.006) (Figure 1, Supplementary Table 7).

**FIGURE 1:**
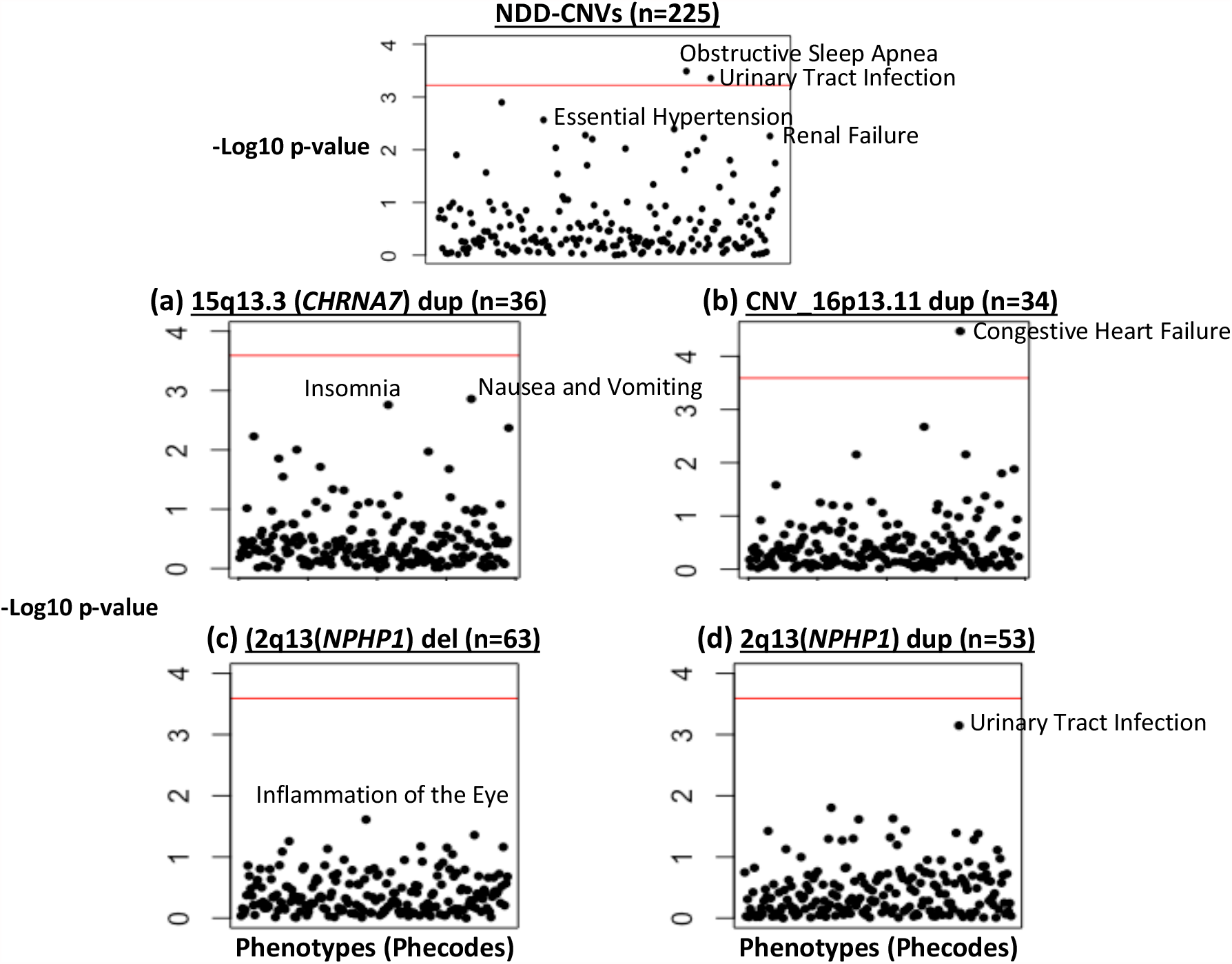
NDD-CNV PheWAS Multi-ancestry, Meta-analysis: Manhattan plots of PheWAS results for multi-ancestry, meta-analysis for: All NDD-CNVs (excluding 15q11.2 del/dup,2q13(NPHP1) del/dup) and most prevalent, individual CNVs (a) 15q13.3 (CHRNA7)dup; (b) 16p13.11dup; (c) 2q13(NPHP1)del and (d) 2q13(NPHP1) dup. *Notes*: -For each Manhattan plot, 195 ICD-based phecodes are on x-axis, y-axis is -log10(p-value) -Red horizontal line represents the Benjamini-Hochberg FDR=0.05 significance threshold -See Supplementary Table 7 and Supplementary Table 8 for detailed PheWAS results, that are depicted in Manhattan plots.

In ancestry-meta-analysis of individual NDD-CNV loci, the 16p13.11 duplication was significant for association with ‘congestive heart failure; non-hypertensive’ (Z-score=4.1, p=3.38×10^−5^) (Figure 1, Supplementary Table 8). In addition, in ancestry-stratified analyses, European-specific PheWAS was significant for association of 15q13.3 duplication (*CHRNA7*) with ‘disorders of vitreous body’ (p=1.02×10^−6^), ‘acute pharyngitis’ (p=1.05×10^−5^), and ‘stiffness of joint’ (p=1.9×10^−4^). Hispanic-specific PheWAS was significant for association of 15q13.3 duplication (*CHRNA7*) and ‘diseases of the larynx and vocal cords’ (p=4.9×10^−7^) and ‘diseases of hair and hair follicles’ (p=4.2×10^−4^). (Supplementary Table 9)

### Association with Clinical Indices (BMI and Lab Values)

Further, we investigated the association of rare NDD-CNVs, in aggregate, with quantitative clinical outcomes, body mass index (BMI) and common serum lab values. The set of all rare, NDD-CNVs (excluding the more common 15q11.2 del/dup and 2q13 NPHP1 del/dup) was found to be associated with increased BMI (n=215 CNV carriers, Beta=0.14, p=0.04) (Supplementary Table 9). Interestingly, as per the ancestry-stratified analyses, for the Hispanic NDD-CNV carriers there was an association of increased BMI with NDD-CNV status (n=74, Beta=0.30, p=0.004), but not for NDD-CNV carriers of African-American (n=75, Beta=0.02, p=0.89) nor European ancestries (n=66, Beta=0.02, p=0.83). Of note, the rare, 16p11.2 deletion (associated with BMI in past reports) was found to be distributed among ancestries (n=3 African-American carriers, n=4 European carrier, n=2 Hispanic carriers).^40^

For serum lab test associations, the aggregated rare, NDD-CNV set was tested for association with 38 common serum lab tests, given their widespread medical utility, including comprehensive metabolic panel, complete blood count and lipid profile tests, that are routinely performed across inpatient and outpatient clinics (Supplementary Table 10). Among the multi-ancestry NDD-CNV carriers, tested in aggregate, there were no significant associations after multiple testing correction.

## DISCUSSION

In the current study, individuals who harbor rare CNVs that are known risk factors for neurodevelopmental disorders, were identified within the Bio*Me* biobank and their clinical presentation surveyed in biobank-based analyses. The leveraged biobank is advantageous in its multi-ancestry composition and robust clinical data repository. Overall, as per the consensus of two CNV calling algorithms, 2.5% of individuals within Bio*Me*, harbor at least one of 64 NDD-CNVs. Ancestry distribution of individuals whom harbored NDD-CNVs was comparable to the ancestry distribution of the biobank, overall (∼1/3 European and ∼1/4 African). Further, 21 NDD-CNVs did not have any carriers within Bio*Me*. The prevalence of individual, rare NDD-CNVs varied in comparison to previous biobank reports, of the UKBB and Geisinger DiscoverEHR, by CNV locus.^19, 20^ The most highly-prevalent NDD-CNVs in Bio*Me*, were also identified as most highly-prevalent in other reports (i.e. 15q11.2deletion/duplication, chr2q13(NPHP1) deletion/duplication). As compared to the Discover EHR cohort, the same NDD-CNVs that had no carriers in DiscoverEHR also had no carriers in Bio*Me*.

In comparing the current analysis of NDD-CNVs in Bio*Me* with previous biobank reports, the divergence in ancestry is most notable, as the UKBB and Geisinger DiscoverEHR cohorts are nearly exclusively of European ancestry (i.e. DiscoverEHR: 98% European ancestry), while Bio*Me* is markedly multi-ancestry. Of further divergence, the UKBB was formed with an intention to recruit a ‘healthy cohort’, in contrast to Bio*Me*, recruiting from within a medical system. Overall, the Bio*Me* biobank is affected by ascertainment bias (i.e., ∼67% recruited from outpatient medicine clinics), as is any biobank, an important caveat in considering biobank comparisons. Within Bio*Me* there is a skew towards older adults with few pediatric participants. The advanced age range presented an opportunity for broad clinical assessment. Indeed, many investigations of NDD-CNVs to date, including their initial discovery, have focused on early-childhood developmental deviation, without consideration of longitudinal illness trajectory. Of note, the healthcare system from which Bio*Me* is derived is not a single-provider, contained system; rather, participants may receive care from other systems. For example, individuals may present to Bio*Me* as an adult, having received care elsewhere as a child/adolescent, or may present for specialty medical care, having received psychiatric care elsewhere. Further, EHR data incorporated in the current analysis dates to 2003-2004, limiting direct, early-life analyses for most adults in the current study.

These challenges of biobank analyses, notwithstanding, the current Bio*Me* analyses yielded interesting and notable findings. Among NDD-CNV carriers, there was a significant enrichment for congenital disorders, confirming previous reports.^5, 6, 19^ Major depressive disorder (MDD) was also found to be significantly enriched, addressing in part, conflicting, past reports about the enrichment of NDD-CNVs in major depressive disorder.^41, 42^ In contrast to the UKBB finding of enrichment of MDD in the cohort of European ancestry, however, the current Bio*Me* analysis yielded enrichment of MDD among individuals of African ancestry, but not Hispanic or European.^41^ The current study used a filtering criteria of at least 2 ICD codes assigned to validate a diagnosis, and a minimum engagement of two clinical encounters, but overall the validity of ICD codes as a proxy for neuropsychiatric diagnosis warrants ongoing assessment in Bio*Me* and other biobanks. Future studies may incorporate phenotyping algorithms, computational tools to mine EHR clinical descriptive data, to identify affected NDD cases, as has been developed for mood disorders.^43^

A previous UKBB analysis tested 58 phenotypes for individual NDD-CNV associations, identifying 46 associations (at FDR threshold of 0.1), including among the most common, obesity, hypertension, and renal failure.^20^ The current Bio*Me* analysis tested an expanded set of 195 phenotypes for association, replicating the UKBB analysis in part, identifying top-most, subthreshold enrichment for hypertension and renal failure for the aggregate, rare NDD-CNV set. Furthermore, the current analysis identified a significant association of the aggregate, rare NDD-CNV set with obstructive sleep apnea (a phenotype not tested in the UKBB analysis), for which obesity is the major risk factor.^44^ The current analysis also identified association of 16p13.11 duplication with congestive heart failure (non-hypertensive), and indeed, case reports of 16p13.11 microduplication indicate increased incidence of congenital heart defects and heart disease.^45^ Other ancestry-specific findings of the 15q13.3 (CHRNA7) duplication, eye disorders in individuals of European ancestry, and disease of the larynx and vocal cords in individuals of Hispanic ancestry, are phenotypes not tested in the previous UKBB analysis, nor have they been widely reported to date in 15q13.3 microduplication in pediatric cases, but may warrant further investigation.^46^

For the aggregate set of NDD-CNVs, there was a significant association with increased BMI, in a multi-ancestry analysis, and further, and an ancestry-specific finding of BMI association among individuals of Hispanic ancestry. While the 16p11.2 deletion is a well-characterized risk factor for obesity, this further implicates the potential role of other NDD-CNVs in obesity.^47^ For 38 common, serum lab tests, including blood count and blood chemistry, there was no identified significant association with the aggregate set of NDD-CNVs.^40^

A limitation of the current analysis is that due to the rarity of NDD-CNVs, the analyses combined NDD-CNVs across loci to ensure well-powered associations, but pooling may dilute the heterogeneity of the effects of individual CNVs. Further, though each CNV was selected based on previous biobank reports as per ‘pathogenicity’ criteria defined by American College of Medical Genetics standards, the strength of evidence varies by NDD-CNV locus.^30, 31^ The associations with quantitative indices were limited to median BMI and serum lab values as outcome measures, but models using generalized linear mixed models may better fit longitudinal biobank data, which can extend over many years with repeated measurements. The current analyses employed select strategies to control for some variables within the EHR (i.e., threshold of number of clinical encounters for inclusion, covarying for density of electronic health records, use of REGENIE method for case/control imbalance), but further methodological innovations and strategies specific to large-scale, biobank analyses may be incorporated as they continue to emerge.^48^

Future recall of individual NDD-CNV carriers may permit detailed clinical measurements beyond EHR-based outcomes, including retrospective, childhood/developmental history, neurocognitive assessments or neuroimaging. The role of other NDD-associated genetic variants may be investigated in future biobank investigations, namely exome sequencing variants (SNVs). In addition, genome-wide CNV burden, the investigation of other genomic CNVs, may further illuminate their role in influencing NDD or NDD-related phenotypes and clinical outcomes. The role of ‘background’ polygenic risk in influencing penetrance, or NDD-case status, is critical, albeit limited in the current multi-ancestry study, as there is a disparity in polygenic risk scores, most derived initially from European cohorts, with trans-ethnic polygenic scores, only recently emerging, and of smaller scale.^49^

Ongoing large-scale, clinical genetic investigations as herein described, based on genetic stratification (i.e., ‘precision psychiatry’), may lead to translational opportunities, clinical insights of at-risk individuals, as well as novel therapeutic strategies targeting specific genetic variants. The importance of diverse inclusion within biobanks and considering the effect of rare, genetic variants in a multi-ancestry context, is evident.

## Data Availability

The output of all data analyses are contained in the Supplementary Tables included in the submission

## ACKNOWLEDGEMENTS

We thank individuals within the BioMe Biobank for their participation. We thank Regeneron Genetics Center for contribution to the exome sequencing and the CLAMMS CNV call set; and the IT group at the Icahn School of Medicine at Mount Sinai for computing and database support.

