## Supplementary Methods and Tables for "A Survey of Copy Number Variants Associated with Neurodevelopmental Disorders in a Large-Scale, Multi-Ancestry Biobank"

*"BioMe" Biobank*

The Bio*Me* Biobank clinical data included in the current analyses consist of longitudinal demographic information, ICD9 and ICD10 codes, laboratory test results, and clinical notes, with clinical data since 2003, increasing in volume and data entry, by progressive year. Participants also provided additional information on self-reported ancestry, personal and family medical history through questionnaires administered upon enrollment. For children under the age of 18, one parent or one legal guardian signed the informed consent form in order for the child (0-18 years of age) to be enrolled. See Supplementary Table 1 for details of recruitment distribution, by clinic site.

*Sample Genotyping and QC*

As previously reported, Bio*Me* biobank participants were genotyped on the Illumina Global Screening Array (GSA) 24v1.0, yielding genotype data for n=32,595 participants at n=635,623 variant sites.[1]

Sample-Level QC: Samples were excluded for gender discordance (genotype sex-check results with homozygosity rates more than 0.2, but less than 0.8 were excluded), low sequencing coverage, heterozygosity rates (samples falling outside of three standard deviations from the mean were excluded), contamination, low call rate, and the discovery of duplicates and sex-discordant samples, yielding n=31,705 samples.

Site-Level QC: Sites were removed with a call rate < 95%, or HWE p-value<1x10-5 (African-American and European ancestry) or p-value<1x10-13 (Hispanic ancestry) yielding 604,869 sites. SHAPEIT/IMPUTE2 were used to pre-phase and impute genotypes using the 1000 Genomes Phase 3 reference panel. Only variants with a minor allele frequency (MAF)>1% and at least a 90% genotyping rate (<10% missing) were included in the final call set for the current analysis.

Relatedness among samples was determined by applying the KING program (Kinship-based Inference for GWAS), which yielded a kinship coefficient for each sample.[2]

### Whole Exome Sequencing and QC

DNA sample preparation and exome sequencing from n=31,249 samples were performed at the Regeneron Genetics Center as previously described.[1] Regions constituting the exome were extracted using the IDT xGen capture platform, and sequenced using 75 bp paired-end sequencing on the Illumina v4 HiSeq 2500 system. Sequence reads were aligned to human reference GRCh37 with BWA-MEM and the resultant BAM files were processed using Picard, and GATK best practice guidelines.[3]

The resulting data was converted into single sample and multi-sample .vcf files, yielding 9,202,884 variants called in 31,250 individuals. Using the Goldilocks Filter (GF), additional filtering was applied[4]: First, a series of filters were applied at particular cells - combinations of site and sample, that is, genotypic information for one individual at one locus. Quality scores were normalized by depth of coverage and used with depth of coverage itself to filter sites, using different thresholds for single nucleotide polymorphisms - SNPs versus small insertions or deletions - Indels. For SNPs, cells with depth-normalized quality scores less than 3, or depth of coverage less than 7 were set to missing. For indels, cells with depth-normalized quality scores less than 5, or depth of coverage less than 10 are set to missing. Then, variant sites were filtered, where all samples carrying variation have heterozygous (0/1) genotype calls, and all samples carrying the heterozygous variation fail the Allele Balance, or AB, cutoff; these sites were removed from the dataset at this stage. The AB cut-off, like the depth and quality scores used for cell filtering above, differed depending on whether the site was a SNP or an indel: SNP variant sites required at least one sample to carry an alternate Allele Balance (AB) >= 15%, and indel variant sites required at least one sample to carry an alternate Allele Balance (AB) >= 20%. These initial site filters resulted in the removal of 441,406 sites, leaving 8,761,478 variants in the Goldilocks-filtered stage.

Sample-Level QC: Post-hoc filtering of the sequenced data including exclusion of low-quality samples including low-coverage, contaminated, and genotype-exome discordant samples; gender discordant and duplicate samples were also removed, yielding n=30,813 samples. Mean depth of coverage for the remaining samples was 36.4x, and a minimum depth of 27.0x, and sequence coverage was sufficient to provide at least 20x haploid read depth at > 85% of targeted bases in 96% of samples.

Site-level QC: Sites with missingness greater than 2% ($n = 267{,}955$ sites), as well as sites showing allele imbalance allelic balance < 0.3 or > 0.8 indicative of a multi-allelic variant ($n = 320{,}877$), yielding 8,172,646 sites.

CNV Calling: WES-based CLAMMS

Sample-Level QC: Samples were excluded with >40 CNV calls or >40,000 exons called as a copy number variant. In addition, for samples with >10% of a chromosome covered by >1 CNV, calls from that chromosome were excluded; the purpose of this filter was to ensure that likely aneuploidies that get fragmented by CLAMMS do not affect the downstream CNV quality control pipeline, particularly for locus definitions. After exclusion of outlier samples, the input for CNV calls was n=26,660 samples.

Site-level QC: A QC score was applied to each CNV call based on call level metrics (Q_nondip & Q_exact), SNP information (het/hom ratio for deletions, het allele balance for duplications), and locus performance over the cohort (e.g. calls heavily enriched in outlier samples flagged as problematic loci). CNV calls with QC scores of 2 and 3 were then retained into a "high-confidence" call set and assigned to a locus definition. CNV super-loci were defined by recursive merging of high-confidence CNVs having >50% reciprocal overlap. Allele frequencies were then calculated on these locus definitions.

CNV Calling: PennCNV

Sample-Level QC: Samples were excluded for waviness factor (WF) >0.05 or <0.05, B Allele Frequency drift>0.01, log R Ratio (LRR) SD>0.3, and numCNV>150, if samples contained more than 150 autosomal CNV calls, yielding a total of **n=30,046 samples**.

Site-level QC: Raw CNV calls were 'merged' so that large CNVs that appear split were combined together if the gap between CNV calls was less than 20%. CNV calls with less than 10 SNP probes were excluded, and CNV calls with >50% overlap of common CNVs cataloged in the database of genomic variants were excluded.

Only Bio*Me* samples with *both* WES-based CLAMMS calls and array-based PennCNV calls, subsequent to QC/filtering (n=24,877 samples) were included in the analysis of NDD-CNV prevalence and downstream phenotypic enrichment and association analyses.

*Filtering for Enrichment and Association analyses*

For the enrichment and association analyses:

Samples with less than two clinical encounters across the EHR were excluded (n=623), to increase the reliability of phenotypic analyses (as less than two clinical encounters likely indicates a lack of engagement in the health system, overall), as well as samples without phenotype data. Furthermore, samples containing the common 15q11.2 del/dup were excluded (n=223), yielding n=22,279 individuals for the analyses of enrichment of neuropsychiatric disorders.

For PheWAS analyses, to reduce potential confounding effects of relatedness, a random individual from each related pair with more than second-degree relatedness (kinship coefficient>0.0885) was excluded (n=1,812), as estimated based on the genotype matrix. In addition, for the ancestry-stratified and multi-ancestry analysis, only individuals of self-reported African, European or Hispanic ancestry were included (excluding others, Native American and Asian), yielding n=19,461 individuals for the PheWAS analyses (n=5,214 African, n=7,082 European, and n=7,165 Hispanic).

*Association with BMI and Serum Lab Values*

For BMI and each of 38 common serum lab tests, a median value across the longitudinal EHR was calculated for each subject. Observations outside of 4 standard deviations from the mean were excluded, suggestive of biologically implausible values due to technical or recording errors, or other extreme variables. Median BMI and serum lab values were inverse normal transformed (INT), as the untransformed measures indicated a non-normal skew in distribution. The resulting transformed values were regressed onto a binary CNV variable and adjusted for covariates of age, sex, ancestry, principal components (PCs) 1-5, and density of records (clinical encounters/years). For each of 38 serum lab tests, individuals with missing outcome measures were excluded, and a multiple testing correction applied.

### SUPPLEMENTARY TABLES

**SUPPLEMENTARY TABLE 1. Bio*Me* Recruitment, by Clinic,** Distribution of Bio*Me* biobank participants by clinic/specialty

**SUPPLEMENTARY TABLE 2. NDD-CNV Prevalence.** Comparison of prevalence of each of 64 NDD-CNVs in Bio*Me* biobank(n=24,877) compared to previously reported UKBB (n=421,268) and Geisinger DiscoverEHR (n=90,595)

**SUPPLEMENTARY TABLE 3: Related NDD-CNV carriers.** Related pairs of individuals in Bio*Me* biobank, both of whom harbor NDD-CNVs (n=17 pairs)

**SUPPLEMENTARY TABLE 4: Enrichment of NDD-CNV carriers for neurodevelopmental and neuropsychiatric disorders, multi-ancestry analyses.** (a) NDD-CNV carriers tested for enrichment, by NDD-CNV (b) Neurodevelopmental and Neuropsychiatric Disorders tested for enrichment, by ICD-10 code

**SUPPLEMENTARY TABLE 5: Enrichment of NDD-CNV carriers, ancestry-stratified.** For two disorders, congenital disorders and major depressive disorder, ancestry-stratified enrichment analysis

**SUPPLEMENTARY TABLE 6: PheWAS of combined NDD-CNVs, multi-ancestry.** PheWAS of an aggregate set of rare, NDD-CNVs, with 195 phenotypes. Association statistics are indicated as well as case-control counts for each phenotype.

**SUPPLEMENTARY TABLE 7: PheWAS of individual NDD-CNVs, multi-ancestry.** PheWAS of four individual, NDD-CNVs with 195 phenotypes. Association statistics are indicated as well as case-control counts for top-most phenotype associations, at $p<0.05$

**SUPPLEMENTARY TABLE 8: PheWAS of individual, prevalent NDD-CNV, ancestry-stratified.** PheWAS of individual, NDD-CNVs, with 195 phenotypes. Association statistics are indicated as well as case-control counts, for top-most associations, at $p<0.05$

**SUPPLEMENTARY TABLE 9: BMI association with combined NDD-CNVs, multi-ancestry and ancestry-stratified.** Association results for BMI with an aggregate set of NDD-CNVs as tabulated by ancestry.

**SUPPLEMENTARY TABLE 10: Serum lab test association with combined NDD-CNVs, multi-ancestry.** Association results for 38 common lab tests with an aggregate set of NDD-CNVs.

**SUPPLEMENTARY TABLE 1. Bio*Me* Recruitment, by Clinic,** Distribution of Bio*Me* biobank participants by clinic/specialty

| Outpatient Clinical Site/Specialty | % |
|---|---|
| Internal Medicine/Primary Care | 66.8 |
| Cadiac Catheterization Laboratory | 8.6 |
| Medical/Surgical | 6.7 |
| Pediatric Allergy | 3.9 |
| Internal Medicine (Pulmonology Focus) | 3.0 |
| HIV/AIDS treatment | 2.7 |
| Surgery/Cardiovascular | 2.7 |
| Movement Disorders | 1.1 |
| OBGYN | 0.8 |
| Cancer Treatment (excluding breast cancer) | 0.8 |
| Multiple Sclerosis | 0.7 |
| General Psychiatry | 0.6 |
| Dermatology | 0.5 |
| Dialysis Center | 0.5 |

| Inpatient Clinical Site/Specialty | % |
|---|---|
| Renal | 0.04 |
| Liver/Hepatic | 0.04 |
| Sleep Medicine | 0.7 |

**SUPPLEMENTARY TABLE 2. NDD-CNV Prevalence.** Comparison of prevalence of each of 64 NDD-CNVs in Bio*Me* biobank(n=24,877) compared to previously reported UKBB (n=421,268) and Geisinger DiscoverEHR (n=90,595)

| | Location (hg19) | Genes (n) | BioMe (n) | Prevalence (%) | Reported UKBB (n) | Prevalence (%) | DiscoverEHR, n | Prevalence (%) |
|---|---|---|---|---|---|---|---|---|
| TAR_del | chr1:145,39–145,81 | 17 | 5 | 0.020 | 75 | 0.018 | | |
| TAR_dup | chr1:145,39–145,81 | 17 | 18 | 0.072 | 436 | 0.100 | | |
| 1q21.1del | chr1:146,53–147,39 | 9 | 8 | 0.032 | 113 | 0.027 | 59 | 0.065 |
| 1q21.1dup | chr1:146,53–147,39 | 9 | 3 | 0.012 | 177 | 0.042 | 90 | 0.099 |
| NRXN1_del | chr2:50,14–51,26 | 1 | 3 | 0.012 | 163 | 0.039 | | |
| 2q11.2del | chr2:96,74–97,68 | 22 | 2 | 0.008 | 31 | 0.007 | | |
| 2q11.2dup | chr2:96,74–97,68 | 22 | 1 | 0.004 | 29 | 0.007 | | |
| 2q13del(NPHP1) | chr2:110,86–110,98 | 3 | 81 | 0.326 | 2448 | 0.580 | | |
| 2q13dup(NPHP1) | chr2:110,86–110,98 | 3 | 62 | 0.249 | 1976 | 0.470 | | |
| 2q13del | chr2:111,39–112,01 | 3 | 3 | 0.012 | 53 | 0.013 | | |
| 2q13dup | chr2:111,39–112,01 | 3 | 8 | 0.032 | 71 | 0.017 | | |
| 2q21.1del | chr2:131,48–131,93 | 5 | 9 | 0.036 | 41 | 0.010 | | |
| 2q21.1dup | chr2:131,48–131,93 | 5 | 5 | 0.020 | 59 | 0.014 | | |
| 3q29del | chr3:195,72–197,35 | 28 | 1 | 0.004 | 9 | 0.002 | 4 | 0.004 |
| 3q29dup | chr3:195,72–197,35 | 28 | 0 | 0 | 5 | 0.001 | | |
| Sotos_5q35del | chr5:175,72-177,05 | 39 | 0 | 0 | 0 | 0 | 0 | 0 |
| 5q35dup | chr5:175,72-177,05 | 39 | 0 | 0 | 0 | 0 | 0 | 0 |
| 7q11.23_del | chr7:72,74–74,14 | 26 | 1 | 0.004 | 1 | 0 | 4 | 0.004 |
| WBS_7q11.23_dup | chr7:72,74–74,14 | 26 | 0 | 0 | 14 | 0.003 | 8 | 0.009 |
| 7q11.23dup_distal | chr7:75,14–76,06 | 16 | 0 | 0 | 24 | 0.006 | | |
| 8p23.1del | chr8:8,10–11,87 | 35 | 0 | 0 | 4 | 0.001 | 0 | 0 |
| 8p23.1dup | chr8:8,10–11,87 | 35 | 0 | 0 | 6 | 0.001 | 0 | 0 |
| 10q11.21q11.23del | chr10:49,39–51,06 | 19 | 1 | 0.004 | 57 | 0.014 | | |
| 10q11.21q11.23dup | chr10:49,39–51,06 | 19 | 2 | 0.008 | 43 | 0.010 | | |
| 10q23del | chr10:82,05–88,93 | 29 | 0 | 0 | 3 | 0.001 | 1 | 0.001 |
| 10q23dup | chr10:82,05–88,93 | 29 | 1 | 0.004 | 7 | 0.002 | | |
| 13q12del(CRYL1) | chr13:20,98–21,10 | 2 | 8 | 0.032 | 379 | 0.090 | | |
| 13q12dup(CRYL1) | chr13:20,98–21,10 | 2 | 0 | 0 | 10 | 0.002 | | |
| 13q12.12del | chr13:23,56–24,88 | 10 | 4 | 0.016 | 85 | 0.020 | | |
| 13q12.12dup | chr13:23,56–24,88 | 10 | 9 | 0.036 | 236 | 0.056 | | |
| 15q11.2del | chr15:22,81–23,09 | 5 | 60 | 0.241 | 1664 | 0.390 | | |
| 15q11.2dup | chr15:22,81–23,09 | 5 | 163 | 0.655 | 2041 | 0.480 | | |
| /AS_15q11.2q13.1 BP1-3_ | chr15:23,68–28,39 | 116 | 0 | 0 | 1 | 0.000 | 5 | 0.006 |
| /AS_15q11.2q13.1 BP1-3_ | chr15:23,68–28,39 | 116 | 0 | 0 | 19 | 0.005 | 3 | 0.003 |
| 11q13del_BP3-BP4(APBA2 | chr15:29,16–30,38 | 4 | 1 | 0.004 | 16 | 0.004 | | |
| 1q13dup_BP3-BP4(APBA2 | chr15:29,16–30,38 | 4 | 3 | 0.012 | 53 | 0.013 | | |
| 15q11q13dup_BP3-BP5 | chr15:29,16–32,46 | 17 | 0 | 0 | 9 | 0.002 | | |
| 15q13.3del | chr15:31,08–32,46 | 8 | 5 | 0.020 | 42 | 0.010 | 55 | 0.061 |
| 15q13.3dup | chr15:31,08–32,46 | 8 | 5 | 0.020 | 240 | 0.057 | | |
| 15q13.3del(CHRNA7) | chr15:32,02–32,46 | 1 | 5 | 0.020 | 10 | 0.002 | | |
| 15q13.3dup(CHRNA7) | chr15:32,02–32,46 | 1 | 42 | 0.169 | 3031 | 0.720 | | |
| 15q24del | chr15:72,90-78,15 | 77 | 0 | 0 | 1 | 0.000 | 2 | 0.002 |
| 15q24dup | chr15:72,90-78,15 | 77 | 0 | 0 | 9 | 0.002 | | |
| 16p13.11del | chr16:15,51–16,29 | 7 | 13 | 0.052 | 131 | 0.031 | 71 | 0.078 |
| 16p13.11dup | chr16:15,51–16,29 | 7 | 39 | 0.157 | 828 | 0.200 | | |
| 16p12.1del | chr16:21,95–22,43 | 8 | 2 | 0.008 | 246 | 0.058 | | |
| 16p12.1dup | chr16:21,95–22,43 | 8 | 14 | 0.056 | 202 | 0.048 | | |
| 16p11.2distal_del | chr16:28,82–29,05 | 11 | 5 | 0.020 | 58 | 0.014 | 28 | 0.031 |
| 16p11.2distal_dup | chr16:28,82–29,05 | 11 | 4 | 0.016 | 137 | 0.033 | | |
| 16p11.2del | chr16:29,65–30,20 | 30 | 15 | 0.060 | 110 | 0.026 | 59 | 0.065 |
| 16p11.2dup | chr16:29,65–30,20 | 30 | 4 | 0.016 | 138 | 0.033 | 63 | 0.07 |
| 17p12del | chr17:14,14–15,43 | 8 | 6 | 0.024 | 237 | 0.056 | 31 | 0.034 |
| 17p12dup | chr17:14,14–15,43 | 8 | 10 | 0.040 | 124 | 0.029 | 38 | 0.042 |
| Magenis Syndrome_17p1 | chr17:16,81–20,21 | 59 | 0 | 0 | 2 | 0.000 | 4 | 0.004 |
| ki-Lupski syndrome_17p11 | chr17:16,81–20,21 | 59 | 0 | 0 | 5 | 0.001 | 0 | 0 |
| 17q11.2del(NF1) | chr17:29,12–30,27 | 19 | 0 | 0 | 9 | 0.002 | 3 | 0.003 |
| 17q11.2dup(NF1) | chr17:29,12–30,27 | 19 | 0 | 0 | 2 | 0.000 | 4 | 0.004 |
| 17q12del | chr17:34,81–36,22 | 17 | 4 | 0.016 | 9 | 0.002 | 4 | 0.004 |
| 17q12dup | chr17:34,81–36,22 | 17 | 4 | 0.016 | 101 | 0.024 | 41 | 0.045 |
| 17q21.31del | chr17:43,70-44,29 | 10 | 0 | 0 | 0 | 0.000 | 0 | 0 |
| 22q11.2del | chr22:19,04–21,47 | 61 | 1 | 0.004 | 10 | 0.002 | 23 | 0.025 |
| 22q11.2dup | chr22:19,04–21,47 | 61 | 7 | 0.028 | 280 | 0.066 | 108 | 0.119 |
| 22q11.2distal_del | chr22:21,92–23,65 | 26 | 0 | 0 | 5 | 0.001 | 1 | 0.001 |
| 22q11.2distal_dup | chr22:21,92–23,65 | 26 | 0 | 0 | 13 | 0.003 | 1 | 0.001 |

*Notes:*

Full References in Main Text: UKBB (*Crawford et al, J Med Genet, 2019* ), Gesinger DiscoverEHR (*Martin et al., JAMA Psychiatry, 2020* )

**SUPPLEMENTARY TABLE 3: Related NDD-CNV carriers.** Related pairs of individuals in Bio*Me* biobank, both of whom harbor NDD-CNVs (n=17 pairs)

| CNV_1 | CNV_2 | Kinship | ANCESTRY |
|---|---|---|---|
| 2q21.1del | 2q21.1del | 0.14 | African American |
| 15q11.2dup | 15q11.2dup | 0.15 | Hispanic |
| 15q11.2del | 15q11.2del | 0.23 | Other |
| 15q13.3del | 15q13.3del | 0.24 | Hispanic |
| 2q13del(NPHP1) | 2q13del(NPHP1) | 0.24 | Other |
| TAR_dup | TAR_dup | 0.25 | Hispanic |
| 2q21.1del | 2q21.1del | 0.25 | African American |
| 2q13dup | 2q13dup | 0.25 | African American |
| 15q11.2dup | 15q11.2dup | 0.25 | African American |
| 15q11.2dup | 15q11.2dup | 0.25 | African American |
| 15q11.2dup | 15q11.2dup | 0.25 | European American |
| 16p13.11dup | 16p13.11dup | 0.25 | African American |
| 2q13del(NPHP1) | 2q13del(NPHP1) | 0.25 | African American |
| 2q21.1del | 2q21.1del | 0.26 | African American |
| TAR_dup | TAR_dup | 0.26 | African American |
| 2q13dup(NPHP1) | 2q13dup(NPHP1) | 0.27 | Hispanic |
| 15q11.2dup | 15q11.2dup | 0.27 | Hispanic |

**SUPPLEMENTARY TABLE 4: Enrichment of NDD-CNV carriers for neurodevelopmental and neuropsychiatric disorders, multi-ancestry analyses.** (a) NDD-CNV carriers tested for enrichment, by NDD-CNV (b) Neurodevelopmental and Neuropsychiatric Disorders tested for enrichment, by ICD-10 code

(a)

| Disorder | ICD Code |
|---|---|
| Schizophrenia and Non-affective Psychosis | F20,F21,F22,F23,F24,F25,F26,F27,F28,F29 |
| ASD and ID | F84, F70,F71,F72,F73,F74,F75,F76,F78,F79 |
| Congenital Disorders | Q0:Q99 |
| Seizure Disorder | G40 |
| Bipolar Disorder | F31 |
| Major Depressive Disorder | F32, F33 |

(b)

| NDD-CNV | COUNT |
|---|---|
| TAR_del | 5 |
| TAR_dup | 15 |
| 1q21.1del | 8 |
| 1q21.1dup | 3 |
| 2q11.2del | 1 |
| 2q11.2dup | 0 |
| 2q13del(NPHP1) | 72 |
| 2q13dup(NPHP1) | 58 |
| 2q13del | 3 |
| 2q13dup | 8 |
| 2q21.1del | 8 |
| 2q21.1dup | 5 |
| 3q29del | 1 |
| 7q11.23_del | 1 |
| 10q11.21q11.23del | 1 |
| 10q11.21q11.23dup | 2 |
| 10q23dup | 1 |
| 13q12del(CRYL1) | 8 |
| 13q12.12del | 3 |
| 13q12.12dup | 9 |
| 15q11q13del_BP3-BP4(APBA2_TJP) | 1 |
| 15q11q13dup_BP3-BP4(APBA2_TJP) | 2 |
| 15q13.3del | 4 |
| 15q13.3dup | 5 |
| 15q13.3del(CHRNA7) | 4 |
| 15q13.3dup(CHRNA7) | 42 |
| 16p13.11del | 12 |
| 16p13.11dup | 39 |
| 16p12.1del | 2 |
| 16p12.1dup | 12 |
| 16p11.2distal_del | 5 |
| 16p11.2distal_dup | 4 |
| 16p11.2del | 12 |
| 16p11.2dup | 4 |
| 17p12del | 4 |
| 17p12dup | 8 |
| 17q12del | 4 |
| 17q12dup | 3 |
| 22q11.2del | 1 |
| 22q11.2dup | 6 |

**SUPPLEMENTARY TABLE 5: Enrichment of NDD-CNV carriers, ancestry-stratified.** For two disorders, congenital disorders and major depressive disorder, ancestry-stratified enrichment analysis

(a)

| CONGENITAL, STRATIFIED | n | p | OR | 95% CI |
|---|---|---|---|---|
| CONGENITAL_AA | 164 | 0.16 | 1.8 | 0.6-4.1 |
| CONGENITAL_EUR | 166 | 1 | 0.8 | 0.1-2.9 |
| CONGENITAL_HISP | 255 | 0.13 | 1.8 | 0.7-3.8 |
| CONGENITAL_OTHER | 54 | 0.005 | 6.9 | 1.7-21.4 |

(b)

| MDD, STRATIFIED | n | p | OR | 95% CI |
|---|---|---|---|---|
| MDD_AA | 907 | 0.01 | 1.8 | 1.1-2.7 |
| MDD_EUR | 793 | 0.54 | 1.2 | 0.6-2.1 |
| MDD_HISP | 1798 | 0.83 | 1.0 | 0.7-1.6 |
| MDD_OTHER | 170 | 0.48 | 1.4 | 0.3-4.7 |

**SUPPLEMENTARY TABLE 6: PheWAS of combined NDD-CNVs, multi-ancestry.** PheWAS of an aggregate set of rare, NDD-CNVs, with 195 phenotypes. Association statistics are indicated as well as case-control counts for each phenotype.

| Phecode | Zscore | p-value | BH | Direction | phenotype | category | AA CASES | AA CONT | EUR CASES | EUR CONT | HISP CASES | HISP CONT |
|---|---|---|---|---|---|---|---|---|---|---|---|---|
| 327.32 | 3.6 | **3.243E-04** | **0.04** | +++ | Obstructive sleep apnea | neurological | 363 | 4719 | 278 | 6686 | 458 | 6539 |
| 591 | 3.5 | **4.402E-04** | **0.04** | +++ | Urinary tract infection | genitourinary | 329 | 4483 | 278 | 6524 | 560 | 6033 |
| 327.3 | 3.2 | 1.27E-03 | 0.08 | +++ | Sleep apnea | neurological | 408 | 4659 | 318 | 6617 | 510 | 6460 |
| 401.1 | 3.0 | 2.72E-03 | 0.13 | +++ | Essential hypertension | circulatory system | 2666 | 2284 | 1650 | 5093 | 3242 | 3560 |
| 401 | 2.9 | 4.08E-03 | 0.14 | +++ | Hypertension | circulatory system | 2673 | 2270 | 1664 | 5068 | 3254 | 3537 |
| 599.3 | 2.8 | 5.30E-03 | 0.14 | +++ | Dysuria | genitourinary | 191 | 4685 | 127 | 6702 | 432 | 6202 |
| 585.1 | 2.8 | 5.52E-03 | 0.14 | +++ | Acute renal failure | genitourinary | 269 | 4722 | 140 | 6817 | 269 | 6667 |
| 428 | 2.7 | 5.97E-03 | 0.14 | +++ | Congestive heart failure; nonhypertensive | circulatory system | 361 | 4708 | 194 | 6746 | 485 | 6493 |
| 278.11 | 2.7 | 6.30E-03 | 0.14 | +++ | Morbid obesity | endocrine/metabolic | 494 | 4463 | 134 | 6862 | 498 | 6395 |
| 465.2 | 2.6 | 9.22E-03 | 0.17 | -++ | Acute pharyngitis | respiratory | 179 | 4688 | 121 | 6623 | 255 | 6349 |
| 327.4 | 2.6 | 0.01 | 0.17 | +++ | Insomnia | neurological | 374 | 4580 | 458 | 6347 | 724 | 6083 |
| 292 | 2.6 | 0.01 | 0.17 | +++ | Neurological disorders | mental disorders | 208 | 4757 | 189 | 6675 | 347 | 6449 |
| 443 | 2.5 | 0.01 | 0.18 | +++ | Peripheral vascular disease | circulatory system | 380 | 4582 | 211 | 6736 | 600 | 6200 |
| 327 | 2.5 | 0.01 | 0.18 | +++ | Sleep disorders | neurological | 437 | 4422 | 551 | 6169 | 823 | 5825 |
| 789 | 2.4 | 0.02 | 0.21 | +++ | Nausea and vomiting | symptoms | 231 | 4578 | 124 | 6706 | 300 | 6319 |
| 532 | 2.4 | 0.02 | 0.22 | +++ | Dysphagia | digestive | 149 | 4912 | 111 | 6857 | 254 | 6690 |
| 443.9 | 2.3 | 0.02 | 0.23 | +++ | Peripheral vascular disease, unspecified | circulatory system | 290 | 4773 | 139 | 6857 | 463 | 6445 |
| 296.22 | 2.3 | 0.02 | 0.26 | +++ | Major depressive disorder | mental disorders | 794 | 4119 | 744 | 6049 | 1500 | 5243 |
| 296.2 | 2.2 | 0.03 | 0.27 | +++ | Depression | mental disorders | 803 | 4100 | 751 | 6038 | 1516 | 5215 |
| 395 | -2.2 | 0.03 | 0.27 | --- | Heart valve disorders | circulatory system | 186 | 4962 | 380 | 6569 | 251 | 6823 |
| 514 | 2.2 | 0.03 | 0.27 | -++ | Abnormal findings examination of lungs | respiratory | 131 | 4990 | 149 | 6813 | 200 | 6850 |
| 296 | 2.0 | 0.05 | 0.40 | +++ | Mood disorders | mental disorders | 873 | 4010 | 841 | 5922 | 1597 | 5110 |
| 585.33 | 1.9 | 0.05 | 0.44 | ++- | Chronic Kidney Disease, Stage III | genitourinary | 436 | 4662 | 263 | 6746 | 555 | 6485 |
| 578.8 | 1.9 | 0.06 | 0.47 | +++ | Hemorrhage of rectum and anus | digestive | 142 | 4851 | 129 | 6727 | 180 | 6692 |
| 611 | 1.8 | 0.07 | 0.54 | ++- | Abnormal findings on mammogram or breast exam | genitourinary | 290 | 4638 | 146 | 6765 | 357 | 6438 |
| 318 | 1.8 | 0.08 | 0.58 | +++ | Tobacco use disorder | mental disorders | 704 | 4266 | 327 | 6604 | 710 | 6157 |
| 287.3 | -1.7 | 0.09 | 0.62 | --- | Thrombocytopenia | hematopoietic | 110 | 5048 | 108 | 6916 | 150 | 6917 |
| 480 | -1.7 | 0.09 | 0.62 | --- | Pneumonia | respiratory | 187 | 4735 | 159 | 6693 | 240 | 6534 |
| 340 | 1.7 | 0.10 | 0.62 | +-+ | Migraine | neurological | 190 | 4875 | 227 | 6719 | 385 | 6533 |
| 721.1 | -1.7 | 0.10 | 0.62 | --- | Spondylosis without myelopathy | musculoskeletal | 240 | 4792 | 217 | 6710 | 367 | 6611 |
| 272.11 | -1.7 | 0.10 | 0.62 | --- | Hypercholesterolemia | endocrine/metabolic | 576 | 4452 | 698 | 6096 | 771 | 6103 |
| 269 | 1.6 | 0.10 | 0.62 | +-+ | Proteinuria | endocrine/metabolic | 262 | 4835 | 164 | 6857 | 360 | 6680 |
| 278.1 | 1.6 | 0.11 | 0.63 | +-+ | Obesity | endocrine/metabolic | 1191 | 3601 | 489 | 6351 | 1357 | 5219 |
| 339 | 1.6 | 0.11 | 0.63 | ++- | Other headache syndromes | neurological | 489 | 4243 | 209 | 6594 | 749 | 5739 |
| 512.9 | 1.6 | 0.11 | 0.63 | +++ | Other dyspnea | respiratory | 508 | 4231 | 436 | 6269 | 670 | 5828 |
| 70 | -1.6 | 0.12 | 0.63 | --- | Viral hepatitis | infectious diseases | 621 | 4513 | 513 | 6522 | 681 | 6399 |
| 365 | 1.5 | 0.12 | 0.63 | ++- | Glaucoma | sense organs | 464 | 4538 | 198 | 6781 | 512 | 6413 |
| 153 | 1.5 | 0.12 | 0.63 | +++ | Colorectal cancer | neoplasms | 112 | 5022 | 149 | 6863 | 129 | 6959 |
| 350.2 | 1.5 | 0.13 | 0.64 | -++ | Abnormality of gait | neurological | 451 | 4547 | 282 | 6620 | 567 | 6289 |
| 418 | 1.5 | 0.13 | 0.64 | +++ | Nonspecific chest pain | circulatory system | 714 | 3763 | 334 | 6215 | 1104 | 4951 |
| 300.1 | 1.5 | 0.14 | 0.64 | +++ | Anxiety disorder | mental disorders | 431 | 4533 | 680 | 6036 | 928 | 5783 |
| 585.34 | 1.5 | 0.14 | 0.64 | ++- | Chronic Kidney Disease, Stage IV | genitourinary | 167 | 5006 | 114 | 6933 | 218 | 6893 |
| 242 | -1.5 | 0.14 | 0.64 | --- | Thyrotoxicosis with or without goiter | endocrine/metabolic | 151 | 5016 | 161 | 6856 | 157 | 6952 |
| 512 | 1.5 | 0.14 | 0.64 | ++- | Other symptoms of respiratory system | respiratory | 1339 | 2892 | 1133 | 5056 | 1886 | 4009 |
| 571.51 | -1.4 | 0.15 | 0.64 | +-- | Cirrhosis of liver without mention of alcohol | digestive | 161 | 4998 | 187 | 6848 | 345 | 6754 |
| 70.3 | -1.4 | 0.15 | 0.65 | --- | Viral hepatitis C | infectious diseases | 530 | 4637 | 445 | 6604 | 628 | 6479 |
| 721 | -1.4 | 0.16 | 0.65 | -+- | Spondylosis and allied disorders | musculoskeletal | 268 | 4754 | 236 | 6684 | 392 | 6571 |
| 599 | 1.4 | 0.16 | 0.65 | -++ | Other symptoms/disorders or the urinary system | genitourinary | 621 | 3843 | 505 | 5921 | 1008 | 5191 |
| 338.2 | 1.4 | 0.16 | 0.66 | +++ | Chronic pain | neurological | 632 | 4165 | 244 | 6616 | 935 | 5698 |
| 338 | 1.3 | 0.19 | 0.71 | +++ | Pain | neurological | 708 | 3948 | 302 | 6378 | 1059 | 5397 |
| 585 | 1.3 | 0.19 | 0.71 | ++- | Renal failure | genitourinary | 883 | 4096 | 494 | 6435 | 936 | 5960 |
| 472 | 1.3 | 0.19 | 0.71 | +++ | Chronic pharyngitis and nasopharyngitis | respiratory | 164 | 4888 | 220 | 6662 | 253 | 6658 |
| 761 | -1.3 | 0.19 | 0.71 | --- | Cervicalgia | symptoms | 283 | 4656 | 186 | 6677 | 446 | 6295 |
| 949 | 1.3 | 0.20 | 0.71 | +-+ | Allergies, other | injuries & poisonings | 140 | 4926 | 123 | 6801 | 277 | 6693 |

**SUPPLEMENTARY TABLE 6: PheWAS of combined NDD-CNVs, multi-ancestry.** PheWAS of an aggregate set of rare, NDD-CNVs, with 195 phenotypes. Association statistics are indicated as well as case-control counts for each phenotype.

| Phecode | Zscore | p-value | BH | Direction | phenotype | category | AA CASES | AA CONT | EUR CASES | EUR CONT | HISP CASES | HISP CONT |
|---|---|---|---|---|---|---|---|---|---|---|---|---|
| 287 | -1.3 | 0.20 | 0.71 | --- | Purpura and other hemorrhagic conditions | hematopoietic | 112 | 5028 | 118 | 6870 | 163 | 6873 |
| 411 | 1.3 | 0.21 | 0.71 | -++ | Ischemic Heart Disease | circulatory system | 510 | 4478 | 611 | 6220 | 1024 | 5824 |
| 745 | 1.3 | 0.21 | 0.71 | -++ | Pain in joint | musculoskeletal | 1328 | 2941 | 687 | 5664 | 1873 | 4121 |
| 763 | -1.2 | 0.22 | 0.73 | -+- | Thoracic or lumbosacral neuritis or radiculitis, unspecified | symptoms | 282 | 4811 | 251 | 6688 | 347 | 6644 |
| 473 | 1.2 | 0.23 | 0.73 | +-+ | Diseases of the larynx and vocal cords | respiratory | 122 | 4973 | 122 | 6816 | 158 | 6872 |
| 351 | -1.2 | 0.23 | 0.73 | --- | Other peripheral nerve disorders | neurological | 188 | 4816 | 130 | 6773 | 263 | 6615 |
| 619 | 1.2 | 0.24 | 0.73 | -++ | Noninflammatory female genital disorders | genitourinary | 290 | 4454 | 102 | 6733 | 426 | 6176 |
| 512.7 | 1.2 | 0.24 | 0.73 | ++- | Shortness of breath | respiratory | 748 | 3921 | 631 | 5939 | 1028 | 5340 |
| 425.1 | -1.2 | 0.24 | 0.73 | --+ | Primary/intrinsic cardiomyopathies | circulatory system | 120 | 5038 | 107 | 6921 | 163 | 6940 |
| 275 | 1.2 | 0.24 | 0.73 | -+- | Disorders of mineral metabolism | endocrine/metabolic | 198 | 4870 | 170 | 6753 | 185 | 6836 |
| 278 | 1.2 | 0.25 | 0.73 | +-+ | Overweight, obesity and other hyperalimentation | endocrine/metabolic | 1422 | 3104 | 735 | 5870 | 1665 | 4605 |
| 585.3 | 1.2 | 0.25 | 0.73 | ++- | Chronic renal failure [CKD] | genitourinary | 734 | 4338 | 436 | 6540 | 820 | 6167 |
| 573.7 | 1.2 | 0.25 | 0.73 | +++ | Abnormal results of function study of liver | digestive | 136 | 4956 | 163 | 6760 | 201 | 6765 |
| 366.2 | 1.1 | 0.26 | 0.75 | +++ | Senile cataract | sense organs | 285 | 4682 | 186 | 6744 | 377 | 6434 |
| 379.2 | 1.1 | 0.27 | 0.76 | ++- | Disorders of vitreous body | sense organs | 126 | 4957 | 108 | 6842 | 151 | 6796 |
| 368 | -1.1 | 0.28 | 0.77 | --- | Visual disturbances | sense organs | 172 | 4608 | 138 | 6708 | 247 | 6235 |
| 78 | -1.1 | 0.28 | 0.77 | -+- | Viral warts & HPV | infectious diseases | 139 | 4933 | 129 | 6820 | 185 | 6781 |
| 350.1 | -1.1 | 0.29 | 0.78 | --+ | Abnormal involuntary movements | neurological | 132 | 4881 | 105 | 6823 | 155 | 6745 |
| 475.9 | 1.0 | 0.30 | 0.78 | -++ | Postnasal drip | respiratory | 113 | 4933 | 112 | 6770 | 112 | 6866 |
| 681 | 1.0 | 0.30 | 0.78 | -++ | Superficial cellulitis and abscess | dermatologic | 235 | 4523 | 167 | 6647 | 384 | 6165 |
| 426 | 1.0 | 0.31 | 0.78 | +-+ | Cardiac conduction disorders | circulatory system | 237 | 4749 | 269 | 6604 | 371 | 6541 |
| 386.9 | 1.0 | 0.32 | 0.78 | +++ | Dizziness and giddiness (Light-headedness and vertigo) | sense organs | 351 | 4482 | 237 | 6514 | 662 | 5893 |
| 241.2 | 1.0 | 0.32 | 0.78 | +-+ | Nontoxic multinodular goiter | endocrine/metabolic | 127 | 5030 | 146 | 6868 | 115 | 6999 |
| 366 | 1.0 | 0.32 | 0.78 | ++- | Cataract | sense organs | 451 | 4415 | 326 | 6503 | 614 | 6069 |
| 300 | 1.0 | 0.32 | 0.78 | -++ | Anxiety disorders | mental disorders | 534 | 4365 | 747 | 5933 | 1078 | 5568 |
| 586 | 1.0 | 0.32 | 0.78 | +-+ | Other disorders of the kidney and ureters | genitourinary | 256 | 4769 | 214 | 6711 | 279 | 6662 |
| 250.4 | 1.0 | 0.33 | 0.78 | -++ | Abnormal glucose | endocrine/metabolic | 285 | 4585 | 263 | 6567 | 335 | 6369 |
| 687.1 | 1.0 | 0.33 | 0.78 | -++ | Rash and other nonspecific skin eruption | dermatologic | 389 | 4351 | 203 | 6518 | 701 | 5678 |
| 389.1 | -1.0 | 0.33 | 0.78 | --+ | Sensorineural hearing loss | sense organs | 105 | 4978 | 128 | 6788 | 232 | 6692 |
| 626 | -1.0 | 0.34 | 0.78 | -+- | Disorders of menstruation and other abnormal bleeding from female genital tract | genitourinary | 361 | 4415 | 208 | 6571 | 453 | 6154 |
| 426.9 | 1.0 | 0.34 | 0.78 | +++ | Cardiac pacemaker/device in situ | circulatory system | 123 | 5073 | 167 | 6858 | 223 | 6904 |
| 357 | 0.9 | 0.35 | 0.78 | ++- | Inflammatory and toxic neuropathy | neurological | 192 | 4868 | 165 | 6816 | 201 | 6800 |
| 760 | 0.9 | 0.35 | 0.78 | ++- | Back pain | symptoms | 1101 | 3467 | 507 | 6105 | 1596 | 4696 |
| 241.1 | -0.9 | 0.35 | 0.78 | --- | Nontoxic uninodular goiter | endocrine/metabolic | 185 | 4944 | 281 | 6684 | 216 | 6865 |
| 578 | 0.9 | 0.36 | 0.78 | -++ | Gastrointestinal hemorrhage | digestive | 226 | 4679 | 185 | 6594 | 303 | 6422 |
| 740.11 | 0.9 | 0.39 | 0.85 | --+ | Osteoarthrosis, localized, primary | musculoskeletal | 526 | 4387 | 351 | 6505 | 604 | 6148 |
| 771 | 0.8 | 0.41 | 0.87 | -++ | Musculoskeletal symptoms referable to limbs | symptoms | 273 | 4804 | 143 | 6867 | 365 | 6645 |
| 429 | 0.8 | 0.44 | 0.87 | +++ | Ill-defined descriptions and complications of heart disease | circulatory system | 126 | 4815 | 108 | 6766 | 199 | 6564 |
| 740.9 | 0.8 | 0.44 | 0.87 | -++ | Osteoarthrosis NOS | musculoskeletal | 492 | 4472 | 246 | 6654 | 842 | 5961 |
| 475 | 0.8 | 0.45 | 0.87 | ++- | Chronic sinusitis | respiratory | 257 | 4648 | 323 | 6400 | 352 | 6446 |
| 452 | -0.8 | 0.45 | 0.87 | +-- | Other venous embolism and thrombosis | circulatory system | 192 | 4935 | 102 | 6932 | 189 | 6881 |
| 250 | 0.7 | 0.46 | 0.87 | +++ | Diabetes mellitus | endocrine/metabolic | 1447 | 3561 | 683 | 6211 | 2026 | 4813 |
| 250.2 | 0.7 | 0.46 | 0.87 | +++ | Type 2 diabetes | endocrine/metabolic | 1438 | 3572 | 612 | 6276 | 2004 | 4831 |
| 939 | -0.7 | 0.48 | 0.87 | -+- | Atopic/contact dermatitis due to other or unspecified | dermatologic | 297 | 4546 | 300 | 6389 | 385 | 6219 |
| 704 | -0.7 | 0.48 | 0.87 | --- | Diseases of hair and hair follicles | dermatologic | 124 | 4839 | 130 | 6754 | 238 | 6540 |
| 530 | 0.7 | 0.48 | 0.87 | -++ | Diseases of esophagus | digestive | 1049 | 3815 | 854 | 5879 | 1819 | 4885 |

**SUPPLEMENTARY TABLE 6: PheWAS of combined NDD-CNVs, multi-ancestry.** PheWAS of an aggregate set of rare, NDD-CNVs, with 195 phenotypes. Association statistics are indicated as well as case-control counts for each phenotype.

| Phecode | Zscore | p-value | BH | Direction | phenotype | category | AA CASES | AA CONT | EUR CASES | EUR CONT | HISP CASES | HISP CONT |
|---|---|---|---|---|---|---|---|---|---|---|---|---|
| 733 | 0.7 | 0.48 | 0.87 | -++ | Other disorders of bone and cartilage | musculoskeletal | 212 | 4866 | 406 | 6486 | 476 | 6474 |
| 455 | 0.7 | 0.48 | 0.87 | -++ | Hemorrhoids | circulatory system | 157 | 4865 | 198 | 6699 | 296 | 6561 |
| 514.2 | 0.7 | 0.49 | 0.87 | +++ | Solitary pulmonary nodule | respiratory | 155 | 4974 | 190 | 6771 | 239 | 6778 |
| 425 | -0.7 | 0.49 | 0.87 | --+ | Cardiomyopathy | circulatory system | 132 | 5022 | 124 | 6902 | 172 | 6925 |
| 627 | -0.7 | 0.50 | 0.87 | +-- | Menopausal and postmenopausal disorders | genitourinary | 243 | 4708 | 205 | 6656 | 312 | 6487 |
| 722 | 0.7 | 0.50 | 0.87 | ++- | Intervertebral disc disorders | musculoskeletal | 184 | 4910 | 179 | 6736 | 226 | 6738 |
| 362 | 0.7 | 0.50 | 0.87 | +++ | Other retinal disorders | sense organs | 231 | 4801 | 185 | 6797 | 270 | 6655 |
| 427.9 | 0.7 | 0.51 | 0.87 | +++ | Palpitations | circulatory system | 245 | 4718 | 225 | 6615 | 432 | 6357 |
| 706 | 0.7 | 0.51 | 0.87 | -++ | Diseases of sebaceous glands | dermatologic | 370 | 4405 | 159 | 6639 | 498 | 6091 |
| 476 | 0.6 | 0.52 | 0.87 | -++ | Allergic rhinitis | respiratory | 635 | 4176 | 569 | 6092 | 952 | 5706 |
| 465 | 0.6 | 0.52 | 0.87 | -++ | Acute upper respiratory infections of multiple or unspecified sites | respiratory | 637 | 3712 | 349 | 6008 | 907 | 5001 |
| 367 | -0.6 | 0.52 | 0.87 | -+- | Disorders of refraction and accommodation; blindness and low vision | sense organs | 142 | 4616 | 106 | 6725 | 242 | 6288 |
| 362.2 | -0.6 | 0.52 | 0.87 | --+ | Degeneration of macula and posterior pole of retina | sense organs | 105 | 5037 | 123 | 6903 | 138 | 6900 |
| 530.1 | 0.6 | 0.52 | 0.87 | -++ | Esophagitis, GERD and related diseases | digestive | 1027 | 3838 | 816 | 5932 | 1758 | 4966 |
| 197 | 0.6 | 0.52 | 0.87 | +++ | Chemotherapy | neoplasms | 206 | 4785 | 205 | 6721 | 235 | 6623 |
| 626.1 | -0.6 | 0.53 | 0.87 | -+- | Irregular menstrual cycle/bleeding | genitourinary | 291 | 4569 | 166 | 6665 | 365 | 6341 |
| 285.2 | -0.6 | 0.53 | 0.87 | +-+ | Anemia of chronic disease | hematopoietic | 136 | 5002 | 103 | 6945 | 124 | 6961 |
| 714 | -0.6 | 0.54 | 0.87 | +-+ | Rheumatoid arthritis and other inflammatory polyarthropathies | musculoskeletal | 140 | 5036 | 109 | 6916 | 205 | 6903 |
| 594 | 0.6 | 0.55 | 0.87 | -++ | Urinary calculus | genitourinary | 104 | 5047 | 184 | 6816 | 258 | 6753 |
| 389 | -0.6 | 0.55 | 0.87 | --+ | Hearing loss | sense organs | 196 | 4728 | 204 | 6578 | 403 | 6325 |
| 740.1 | 0.6 | 0.55 | 0.87 | --+ | Osteoarthritis; localized | musculoskeletal | 542 | 4362 | 365 | 6490 | 632 | 6105 |
| 782.3 | 0.6 | 0.55 | 0.87 | -++ | Edema | symptoms | 381 | 4455 | 211 | 6613 | 448 | 6263 |
| 379 | 0.6 | 0.56 | 0.87 | ++- | Other disorders of eye | sense organs | 140 | 4896 | 124 | 6808 | 175 | 6719 |
| 375.1 | 0.6 | 0.57 | 0.87 | -++ | Dry eyes | sense organs | 228 | 4725 | 178 | 6724 | 435 | 6323 |
| 765 | -0.6 | 0.57 | 0.87 | --- | Cervical radiculitis | symptoms | 131 | 4995 | 101 | 6898 | 177 | 6877 |
| 571.5 | -0.6 | 0.57 | 0.87 | +-- | Other chronic nonalcoholic liver disease | digestive | 247 | 4867 | 324 | 6632 | 514 | 6489 |
| 798 | -0.6 | 0.57 | 0.87 | -+- | Malaise and fatigue | symptoms | 587 | 3922 | 574 | 5782 | 794 | 5426 |
| 550 | 0.6 | 0.57 | 0.87 | ++- | Abdominal hernia | digestive | 277 | 4827 | 252 | 6714 | 399 | 6598 |
| 593 | 0.5 | 0.59 | 0.88 | --+ | Hematuria | genitourinary | 151 | 4883 | 129 | 6778 | 234 | 6671 |
| 740 | 0.5 | 0.59 | 0.88 | +++ | Osteoarthrosis | musculoskeletal | 852 | 3979 | 548 | 6194 | 1229 | 5422 |
| 571 | -0.5 | 0.59 | 0.88 | +-- | Chronic liver disease and cirrhosis | digestive | 255 | 4857 | 359 | 6594 | 547 | 6453 |
| 479 | -0.5 | 0.60 | 0.88 | --- | Other upper respiratory disease | respiratory | 193 | 4734 | 137 | 6698 | 245 | 6511 |
| 174.11 | 0.5 | 0.60 | 0.88 | +-+ | Malignant neoplasm of female breast | neoplasms | 152 | 5033 | 262 | 6776 | 182 | 6949 |
| 726 | -0.5 | 0.61 | 0.88 | --+ | Peripheral enthesopathies and allied syndromes | musculoskeletal | 352 | 4461 | 275 | 6489 | 486 | 6058 |
| 427.2 | -0.5 | 0.61 | 0.88 | +-- | Atrial fibrillation and flutter | circulatory system | 194 | 4959 | 388 | 6560 | 294 | 6788 |
| 285 | 0.5 | 0.62 | 0.88 | ++- | Other anemias | hematopoietic | 1067 | 3604 | 528 | 6161 | 1114 | 5412 |
| 411.4 | 0.5 | 0.62 | 0.88 | -+- | Coronary atherosclerosis | circulatory system | 460 | 4621 | 588 | 6271 | 933 | 6045 |
| 512.8 | 0.5 | 0.62 | 0.88 | -++ | Cough | respiratory | 675 | 3891 | 509 | 6032 | 964 | 5412 |
| 565 | -0.5 | 0.63 | 0.88 | --+ | Anal and rectal conditions | digestive | 146 | 4944 | 187 | 6735 | 188 | 6806 |
| 244 | -0.5 | 0.63 | 0.88 | -++ | Hypothyroidism | endocrine/metabolic | 292 | 4831 | 924 | 5902 | 701 | 6326 |
| 90 | -0.5 | 0.63 | 0.88 | +-- | Sexually transmitted infections (not HIV or hepatitis) | infectious diseases | 102 | 4985 | 145 | 6851 | 115 | 6939 |
| 580 | 0.5 | 0.64 | 0.88 | --+ | Nephritis; nephrosis; renal sclerosis | genitourinary | 130 | 5021 | 118 | 6924 | 145 | 6940 |
| 350 | 0.5 | 0.64 | 0.88 | -++ | Abnormal movement | neurological | 555 | 4296 | 362 | 6417 | 693 | 5982 |
| 741.2 | 0.4 | 0.66 | 0.89 | -++ | Stiffness of joint | musculoskeletal | 146 | 5022 | 111 | 6933 | 202 | 6909 |
| 261.2 | 0.4 | 0.66 | 0.89 | +-+ | Vitamin B-complex deficiencies | endocrine/metabolic | 214 | 4917 | 220 | 6737 | 327 | 6705 |
| 530.11 | 0.4 | 0.67 | 0.89 | -++ | GERD | digestive | 1009 | 3869 | 745 | 6022 | 1717 | 5030 |
| 54 | 0.4 | 0.68 | 0.89 | -++ | Herpes simplex | infectious diseases | 159 | 4953 | 169 | 6797 | 178 | 6828 |
| 208 | -0.4 | 0.68 | 0.89 | -+- | Benign neoplasm of colon | neoplasms | 187 | 4849 | 159 | 6757 | 241 | 6683 |
| 271 | -0.4 | 0.69 | 0.89 | --+ | Disorders of carbohydrate transport and metabolism | endocrine/metabolic | 657 | 4276 | 279 | 6638 | 757 | 6060 |
| 272 | 0.4 | 0.69 | 0.89 | --+ | Disorders of lipoid metabolism | endocrine/metabolic | 1736 | 3136 | 1828 | 4731 | 2655 | 4012 |

**SUPPLEMENTARY TABLE 6: PheWAS of combined NDD-CNVs, multi-ancestry.** PheWAS of an aggregate set of rare, NDD-CNVs, with 195 phenotypes. Association statistics are indicated as well as case-control counts for each phenotype.

| Phecode | Zscore | p-value | BH | Direction | phenotype | category | AA CASES | AA CONT | EUR CASES | EUR CONT | HISP CASES | HISP CONT |
|---|---|---|---|---|---|---|---|---|---|---|---|---|
| 271.3 | -0.4 | 0.69 | 0.90 | --+ | Intestinal disaccharidase deficiencies and disaccharide malabsorption | endocrine/metabolic | 655 | 4283 | 274 | 6646 | 756 | 6064 |
| 71 | -0.4 | 0.71 | 0.91 | -+- | Human immunodeficiency virus [HIV] disease | infectious diseases | 429 | 4737 | 361 | 6705 | 421 | 6686 |
| 427 | 0.4 | 0.72 | 0.91 | +-+ | Cardiac dysrhythmias | circulatory system | 588 | 4143 | 713 | 5907 | 913 | 5632 |
| 785 | 0.3 | 0.73 | 0.91 | +-+ | Abdominal pain | symptoms | 771 | 3453 | 463 | 5899 | 1379 | 4449 |
| 276.1 | 0.3 | 0.74 | 0.91 | +-- | Electrolyte imbalance | endocrine/metabolic | 420 | 4488 | 285 | 6598 | 527 | 6294 |
| 272.13 | -0.3 | 0.74 | 0.91 | ++- | Mixed hyperlipidemia | endocrine/metabolic | 197 | 4909 | 227 | 6718 | 308 | 6707 |
| 274 | 0.3 | 0.74 | 0.91 | -+- | Gout and other crystal arthropathies | endocrine/metabolic | 159 | 4999 | 147 | 6873 | 151 | 6964 |
| 71.1 | -0.3 | 0.75 | 0.91 | ++- | HIV infection, symptomatic | infectious diseases | 406 | 4755 | 332 | 6726 | 395 | 6711 |
| 371 | 0.3 | 0.75 | 0.91 | ++- | Inflammation of the eye | sense organs | 131 | 4888 | 148 | 6732 | 168 | 6675 |
| 250.3 | -0.3 | 0.76 | 0.91 | --+ | Insulin pump user | endocrine/metabolic | 288 | 4857 | 149 | 6895 | 432 | 6605 |
| 280 | 0.3 | 0.76 | 0.91 | -+- | Iron deficiency anemias | hematopoietic | 432 | 4516 | 184 | 6727 | 492 | 6376 |
| 244.4 | 0.3 | 0.76 | 0.91 | -++ | Hypothyroidism NOS | endocrine/metabolic | 240 | 4897 | 699 | 6156 | 610 | 6422 |
| 252.1 | 0.3 | 0.77 | 0.91 | ++- | Hyperparathyroidism | endocrine/metabolic | 114 | 5061 | 102 | 6937 | 102 | 7031 |
| 274.1 | -0.3 | 0.77 | 0.91 | -+- | Gout | endocrine/metabolic | 151 | 5010 | 138 | 6890 | 140 | 6981 |
| 600 | -0.3 | 0.79 | 0.93 | --- | Hyperplasia of prostate | genitourinary | 219 | 4888 | 328 | 6593 | 342 | 6711 |
| 495 | 0.3 | 0.80 | 0.93 | +-+ | Asthma | respiratory | 773 | 4233 | 523 | 6322 | 1277 | 5576 |
| 433 | 0.3 | 0.80 | 0.93 | +-+ | Cerebrovascular disease | circulatory system | 359 | 4712 | 207 | 6663 | 436 | 6533 |
| 599.5 | -0.2 | 0.81 | 0.94 | -+- | Frequency of urination and polyuria | genitourinary | 276 | 4515 | 292 | 6428 | 390 | 6230 |
| 280.1 | 0.2 | 0.84 | 0.97 | -+- | Iron deficiency anemias, unspecified or not due to blood loss | hematopoietic | 359 | 4652 | 152 | 6818 | 428 | 6508 |
| 279.7 | -0.2 | 0.84 | 0.97 | -+- | Other immunological findings | endocrine/metabolic | 275 | 4781 | 217 | 6749 | 345 | 6631 |
| 726.1 | 0.2 | 0.85 | 0.97 | --+ | Enthesopathy | musculoskeletal | 160 | 4804 | 139 | 6759 | 244 | 6544 |
| 790.6 | -0.2 | 0.87 | 0.97 | --+ | Other abnormal blood chemistry | symptoms | 281 | 4577 | 265 | 6510 | 270 | 6522 |
| 272.1 | 0.2 | 0.87 | 0.97 | --+ | Hyperlipidemia | endocrine/metabolic | 1723 | 3150 | 1786 | 4784 | 2629 | 4043 |
| 241 | -0.2 | 0.87 | 0.97 | +-- | Nontoxic nodular goiter | endocrine/metabolic | 319 | 4755 | 430 | 6476 | 319 | 6727 |
| 605 | 0.2 | 0.87 | 0.97 | -+- | Erectile dysfunction [ED] | genitourinary | 313 | 4772 | 263 | 6670 | 280 | 6726 |
| 261 | -0.2 | 0.87 | 0.97 | +-- | Vitamin deficiency | endocrine/metabolic | 1288 | 3501 | 1338 | 5202 | 1688 | 4980 |
| 174.1 | 0.1 | 0.88 | 0.97 | --+ | Breast cancer [female] | neoplasms | 176 | 5017 | 284 | 6747 | 199 | 6936 |
| 110.1 | -0.1 | 0.88 | 0.97 | --+ | Dermatophytosis | infectious diseases | 419 | 4399 | 136 | 6738 | 589 | 6041 |
| 276.13 | -0.1 | 0.89 | 0.97 | ++- | Hyperpotassemia | endocrine/metabolic | 197 | 4866 | 174 | 6797 | 298 | 6675 |
| 695 | -0.1 | 0.90 | 0.97 | -+- | Erythematous conditions | dermatologic | 200 | 4880 | 144 | 6803 | 300 | 6618 |
| 687.4 | -0.1 | 0.91 | 0.97 | --+ | Disturbance of skin sensation | dermatologic | 232 | 4590 | 132 | 6691 | 270 | 6385 |
| 573.9 | 0.1 | 0.91 | 0.97 | +-+ | Abnormal serum enzyme levels | digestive | 124 | 4948 | 113 | 6846 | 147 | 6840 |
| 174 | 0.1 | 0.91 | 0.97 | --+ | Breast cancer | neoplasms | 179 | 5007 | 288 | 6740 | 202 | 6923 |
| 727 | 0.1 | 0.93 | 0.98 | -++ | Other disorders of synovium, tendon, and bursa | musculoskeletal | 179 | 4803 | 108 | 6812 | 266 | 6576 |
| 261.4 | -0.1 | 0.93 | 0.98 | +-- | Vitamin D deficiency | endocrine/metabolic | 1195 | 3613 | 1255 | 5317 | 1542 | 5163 |
| 496 | -0.1 | 0.93 | 0.98 | -+- | Chronic airway obstruction | respiratory | 307 | 4780 | 281 | 6690 | 422 | 6593 |
| 743.1 | -0.1 | 0.94 | 0.98 | -+- | Osteoporosis | musculoskeletal | 257 | 4688 | 464 | 6264 | 557 | 6230 |
| 110 | 0.1 | 0.95 | 0.98 | -++ | Dermatophytosis / Dermatomycosis | infectious diseases | 461 | 4313 | 153 | 6692 | 627 | 5928 |
| 427.21 | -0.1 | 0.95 | 0.98 | +-- | Atrial fibrillation | circulatory system | 175 | 4976 | 371 | 6585 | 272 | 6810 |
| 276 | 0.1 | 0.96 | 0.98 | +-- | Disorders of fluid, electrolyte, and acid-base balance | endocrine/metabolic | 446 | 4365 | 313 | 6488 | 553 | 6169 |
| 741 | -0.1 | 0.96 | 0.98 | -+- | Symptoms and disorders of the joints | musculoskeletal | 229 | 4748 | 156 | 6764 | 290 | 6599 |
| 252 | 0.0 | 0.97 | 0.98 | +-- | Disorders of parathyroid gland | endocrine/metabolic | 131 | 5040 | 119 | 6913 | 117 | 7010 |
| 743 | 0.0 | 0.97 | 0.98 | -+- | Osteoporosis, osteopenia and pathological fracture | musculoskeletal | 288 | 4627 | 484 | 6191 | 585 | 6170 |
| 250.42 | 0.0 | 0.98 | 0.99 | -++ | Other abnormal glucose | endocrine/metabolic | 226 | 4672 | 209 | 6652 | 269 | 6468 |
| 743.11 | 0.0 | 1.00 | 1.00 | -+- | Osteoporosis NOS | musculoskeletal | 252 | 4719 | 461 | 6292 | 550 | 6265 |

**SUPPLEMENTARY TABLE 7: PheWAS of individual NDD-CNVs, multi-ancestry.** PheWAS of four individual, NDD-CNVs with 195 phenotypes. Association statistics are indicated as well as case-control counts for top-most phenotype associations, at $p<0.05$

| NDD CNV | Phecode | Zscore | P.value | BH | Direction | phenotype | category | AA CASES | AA CONT | EUR CASES | EUR CONT | HISP CASES | HISP CONT |
|---|---|---|---|---|---|---|---|---|---|---|---|---|---|
| 15q13.3dup_CHRNA7 | 789 | 3.2 | 0.001 | 0.17 | +++ | Nausea and vomiting | symptoms | 231 | 4578 | 124 | 6706 | 300 | 6319 |
| 15q13.3dup_CHRNA7 | 327.4 | 3.1 | 0.002 | 0.17 | +++ | Insomnia | neurological | 374 | 4580 | 458 | 6347 | 724 | 6083 |
| 15q13.3dup_CHRNA7 | 578.8 | 2.9 | 0.004 | 0.28 | ++- | Hemorrhage of rectum and anus | digestive | 142 | 4851 | 129 | 6727 | 180 | 6692 |
| 15q13.3dup_CHRNA7 | 327 | 2.8 | 0.01 | 0.29 | +++ | Sleep disorders | neurological | 437 | 4422 | 551 | 6169 | 823 | 5825 |
| 15q13.3dup_CHRNA7 | 379.2 | 2.6 | 0.01 | 0.35 | -+- | Disorders of vitreous body | sense organs | 126 | 4957 | 108 | 6842 | 151 | 6796 |
| 15q13.3dup_CHRNA7 | 473 | 2.6 | 0.01 | 0.35 | --+ | Diseases of the larynx and vocal cords | respiratory | 122 | 4973 | 122 | 6816 | 158 | 6872 |
| 15q13.3dup_CHRNA7 | 578 | 2.5 | 0.01 | 0.39 | ++- | Gastrointestinal hemorrhage | digestive | 226 | 4679 | 185 | 6594 | 303 | 6422 |
| 15q13.3dup_CHRNA7 | 300 | 2.3 | 0.02 | 0.46 | +++ | Anxiety disorders | mental disorders | 534 | 4365 | 747 | 5933 | 1078 | 5568 |
| 15q13.3dup_CHRNA7 | 350.2 | 2.3 | 0.02 | 0.46 | +++ | Abnormality of gait | neurological | 451 | 4547 | 282 | 6620 | 567 | 6289 |
| 15q13.3dup_CHRNA7 | 300.1 | 2.2 | 0.03 | 0.55 | +++ | Anxiety disorder | mental disorders | 431 | 4533 | 680 | 6036 | 928 | 5783 |
| 15q13.3dup_CHRNA7 | 465.2 | 2.0 | 0.05 | 0.78 | -+- | Acute pharyngitis | respiratory | 179 | 4688 | 121 | 6623 | 255 | 6349 |
| 15q13.3dup_CHRNA7 | 681 | 2.0 | 0.05 | 0.78 | +++ | Superficial cellulitis and abscess | dermatologic | 235 | 4523 | 167 | 6647 | 384 | 6165 |
| 16p13.11_dup | 428 | 4.1 | 3.38E-05 | 0.01 | +++ | Congestive heart failure; nonhypertensive | circulatory system | 361 | 4708 | 194 | 6746 | 485 | 6493 |
| 16p13.11_dup | 241 | 3.1 | 0.002 | 0.21 | +++ | Nontoxic nodular goiter | endocrine/metabolic | 319 | 4755 | 430 | 6476 | 319 | 6727 |
| 16p13.11_dup | 591 | 2.7 | 0.007 | 0.34 | +++ | Urinary tract infection | genitourinary | 329 | 4483 | 278 | 6524 | 560 | 6033 |
| 16p13.11_dup | 733 | 2.7 | 0.01 | 0.34 | -++ | Other disorders of bone and cartilage | musculoskeletal | 212 | 4866 | 406 | 6486 | 476 | 6474 |
| 16p13.11_dup | 512 | 2.5 | 0.01 | 0.51 | +++ | Other symptoms of respiratory system | respiratory | 1339 | 2892 | 1133 | 5056 | 1886 | 4009 |
| 16p13.11_dup | 949 | 2.4 | 0.02 | 0.52 | ++- | Allergies, other | injuries & poisonings | 140 | 4926 | 123 | 6801 | 277 | 6693 |
| 16p13.11_dup | 278 | 2.2 | 0.03 | 0.73 | +++ | Overweight, obesity and other hyperalimentation | endocrine/metabolic | 1422 | 3104 | 735 | 5870 | 1665 | 4605 |
| 16p13.11_dup | 285 | 2.0 | 0.04 | 0.86 | ++- | Other anemias | hematopoietic | 1067 | 3604 | 528 | 6161 | 1114 | 5412 |
| 16p13.11_dup | 241.2 | 2.0 | 0.05 | 0.86 | +-+ | Nontoxic multinodular goiter | endocrine/metabolic | 127 | 5030 | 146 | 6868 | 115 | 6999 |
| 16p13.11_dup | 278.11 | 1.9 | 0.05 | 0.86 | +++ | Morbid obesity | endocrine/metabolic | 494 | 4463 | 134 | 6862 | 498 | 6395 |
| CNV_NPHP1_DEL | 371 | 2.2 | 0.02 | 0.94 | +-+ | Inflammation of the eye | sense organs | 131 | 4888 | 148 | 6732 | 168 | 6675 |
| CNV_NPHP1_DEL | 285 | -2.0 | 0.04 | 0.94 | --- | Other anemias | hematopoietic | 1067 | 3604 | 528 | 6161 | 1114 | 5412 |
| CNV_NPHP1_DUP | 591 | 3.4 | 0.001 | 0.14 | -++ | Urinary tract infection | genitourinary | 329 | 4483 | 278 | 6524 | 560 | 6033 |
| CNV_NPHP1_DUP | 530.11 | 2.4 | 0.02 | 0.81 | +++ | GERD | digestive | 1009 | 3869 | 745 | 6022 | 1717 | 5030 |
| CNV_NPHP1_DUP | 272.11 | -2.3 | 0.02 | 0.81 | --- | Hypercholesterolemia | endocrine/metabolic | 576 | 4452 | 698 | 6096 | 771 | 6103 |
| CNV_NPHP1_DUP | 276 | -2.3 | 0.02 | 0.81 | --- | Disorders of fluid, electrolyte, and acid-base balance | endocrine/metabolic | 446 | 4365 | 313 | 6488 | 553 | 6169 |
| CNV_NPHP1_DUP | 110.1 | 2.1 | 0.04 | 0.81 | -++ | Dermatophytosis | infectious diseases | 419 | 4399 | 136 | 6738 | 589 | 6041 |
| CNV_NPHP1_DUP | 276.1 | -2.1 | 0.04 | 0.81 | --- | Electrolyte imbalance | endocrine/metabolic | 420 | 4488 | 285 | 6598 | 527 | 6294 |
| CNV_NPHP1_DUP | 530 | 2.0 | 0.04 | 0.81 | +++ | Diseases of esophagus | digestive | 1049 | 3815 | 854 | 5879 | 1819 | 4885 |
| CNV_NPHP1_DUP | 285 | 2.0 | 0.04 | 0.81 | +++ | Other anemias | hematopoietic | 1067 | 3604 | 528 | 6161 | 1114 | 5412 |
| CNV_NPHP1_DUP | 110 | 2.0 | 0.05 | 0.81 | -++ | Dermatophytosis / Dermatomycosis | infectious diseases | 461 | 4313 | 153 | 6692 | 627 | 5928 |
| CNV_NPHP1_DUP | 580 | 2.0 | 0.05 | 0.81 | -++ | Nephritis; nephrosis; renal sclerosis | genitourinary | 130 | 5021 | 118 | 6924 | 145 | 6940 |
| CNV_NPHP1_DUP | 530.1 | 2.0 | 0.05 | 0.81 | +++ | Esophagitis, GERD and related diseases | digestive | 1027 | 3838 | 816 | 5932 | 1758 | 4966 |

**<u>SUPPLEMENTARY TABLE 8</u>: PheWAS of individual, prevalent NDD-CNV, ancestry-stratified.** PheWAS of individual, NDD-CNVs, with 195 phenotypes. Association statistics are indicated as well as case-control counts, for top-most associations, at p<0.05

| Ancestry | CNV | phecodes | BETA | SE | CHISQ | p | FDR_BH | phenotype | category | CASES | CONT |
|---|---|---|---|---|---|---|---|---|---|---|---|
| EUR | 15q13.3dup_CHRNA7 | 379.2 | 8.90 | 1.82 | 23.9 | 1.02E-06 | 2.00E-04 | Disorders of vitreous body | sense organs | 108 | 6842 |
| EUR | 15q13.3dup_CHRNA7 | 465.2 | 7.41 | 1.68 | 19.4 | 1.05E-05 | 1.02E-03 | Acute pharyngitis | respiratory | 121 | 6623 |
| EUR | 15q13.3dup_CHRNA7 | 379 | 5.99 | 1.55 | 15.0 | 1.06E-04 | 0.007 | Other disorders of eye | sense organs | 124 | 6808 |
| EUR | 15q13.3dup_CHRNA7 | 741.2 | 5.59 | 1.50 | 13.9 | 1.90E-04 | 0.009 | Stiffness of joint | musculoskeletal | 111 | 6933 |
| EUR | 15q13.3dup_CHRNA7 | 578.8 | 6.27 | 1.91 | 10.8 | 0.001 | 0.04 | Hemorrhage of rectum and anus | digestive | 129 | 6727 |
| EUR | 15q13.3dup_CHRNA7 | 578 | 5.88 | 1.86 | 10.0 | 0.002 | 0.05 | Gastrointestinal hemorrhage | digestive | 185 | 6594 |
| EUR | 16p13.11dup | 428 | 9.72 | 2.32 | 17.54 | 2.82E-05 | 0.005 | Congestive heart failure; nonhypertensive | circulatory system | 194 | 6746 |
| EUR | 16p13.11dup | 733 | 8.82 | 2.22 | 15.74 | 7.28E-05 | 0.007 | Other disorders of bone and cartilage | musculoskeletal | 406 | 6486 |
| EUR | NPHP1_DUP | 591 | 7.29 | 1.67 | 19.10 | 1.24E-05 | 2.42E-03 | Urinary tract infection | genitourinary | 278 | 6524 |
| HISP | 15q13.3dup_CHRNA7 | 473 | 13.60 | 2.70 | 25.30 | 4.90E-07 | 9.56E-05 | Diseases of the larynx and vocal cords | respiratory | 122 | 6816 |
| HISP | 15q13.3dup_CHRNA7 | 704 | 7.15 | 2.03 | 12.44 | 4.21E-04 | 4.10E-02 | Diseases of hair and hair follicles | dermatologic | 130 | 6754 |

**SUPPLEMENTARY TABLE 9: BMI association with combined NDD-CNVs, multi-ancestry and ancestry-stratified.** Association results for BMI with an aggregate set of NDD-CNVs as tabulated by ancestry.

(a)

| NDD-CNV | AA COUNT | EUR COUNT | HISP COUNT |
|---|---|---|---|
| TAR_del | 3 | 0 | 2 |
| TAR_dup | 4 | 4 | 4 |
| 1q21.1del | 2 | 1 | 4 |
| 1q21.1dup | 0 | 2 | 1 |
| NRXN1_DEL | 0 | 1 | 2 |
| 2q11.2del | 0 | 1 | 1 |
| 2q11.2dup | 1 | 0 | 0 |
| 2q13del | 0 | 1 | 1 |
| 2q13dup | 5 | 2 | 1 |
| 2q21.1del | 3 | 1 | 2 |
| 2q21.1dup | 4 | 1 | 0 |
| 7q11.23_del | 0 | 0 | 1 |
| 10q11.21q11.23del | 0 | 1 | 0 |
| 10q11.21q11.23dup | 1 | 1 | 0 |
| 10q23dup | 0 | 0 | 1 |
| 13q12.12del | 2 | 0 | 1 |
| 13q12.12dup | 2 | 2 | 3 |
| 13q12del(CRYL1) | 0 | 7 | 1 |
| 15q11q13del_BP3-BP4(APBA2_TJP) | 0 | 0 | 1 |
| 15q11q13dup_BP3-BP4(APBA2_TJP) | 1 | 0 | 1 |
| 15q13.3del | 2 | 0 | 1 |
| 15q13.3del(CHRNA7) | 2 | 0 | 1 |
| 15q13.3dup | 2 | 1 | 1 |
| 15q13.3dup(CHRNA7) | 7 | 15 | 9 |
| 16p11.2del | 3 | 4 | 2 |
| 16p11.2distal_del | 2 | 0 | 0 |
| 16p11.2distal_dup | 2 | 1 | 0 |
| 16p11.2dup | 0 | 3 | 1 |
| 16p12.1del | 0 | 2 | 0 |
| 16p12.1dup | 3 | 1 | 6 |
| 16p13.11del | 1 | 5 | 6 |
| 16p13.11dup | 14 | 7 | 13 |
| 17p12del | 1 | 2 | 1 |
| 17p12dup | 3 | 0 | 4 |
| 17q12del | 3 | 0 | 1 |
| 17q12dup | 2 | 1 | 1 |
| 22q11.2del | 0 | 0 | 1 |
| 22q11.2dup | 4 | 0 | 1 |

(b)

| | p_value | Estimate | SE |
|---|---|---|---|
| MULTI-ANCESTRY | 0.04 | 0.14 | 2.09 |
| AA STRATIFIED | 0.89 | 0.02 | 0.13 |
| EUR STRATIFIED | 0.83 | 0.02 | 0.21 |
| HISP STRATIFIED | 4.24E-03 | 0.30 | 2.86 |

**SUPPLEMENTARY TABLE 10: Serum lab test association with combined NDD-CNVs, multi-ancestry.**
Association results for 38 common lab tests with an aggregate set of NDD-CNVs.

(a)

| NDD-CNV | COUNT (Multi-ancestry) |
|---|---|
| TAR_del | 3 |
| TAR_dup | 12 |
| NRXN1_DEL | 2 |
| 2q11.2del | 1 |
| 2q11.2dup | 1 |
| 2q13del | 3 |
| 2q13del(NPHP1) | 0 |
| 2q13dup | 7 |
| 2q21.1del | 7 |
| 2q21.1dup | 5 |
| 7q11.23_del | 1 |
| 10q11.21q11.23del | 1 |
| 10q11.21q11.23dup | 2 |
| 10q23dup | 1 |
| 13q12.12del | 4 |
| 13q12.12dup | 8 |
| 13q12del(CRYL1) | 7 |
| 15q11q13del_BP3-BP4(APBA2_TJP) | 1 |
| 15q11q13dup_BP3-BP4(APBA2_TJP) | 2 |
| 15q13.3del | 3 |
| 15q13.3del(CHRNA7) | 3 |
| 15q13.3dup | 5 |
| 15q13.3dup(CHRNA7) | 40 |
| 16p11.2del | 10 |
| 16p11.2distal_del | 1 |
| 16p11.2distal_dup | 4 |
| 16p11.2dup | 4 |
| 16p12.1del | 2 |
| 16p12.1dup | 11 |
| 16p13.11del | 12 |
| 16p13.11dup | 33 |
| 17p12del | 4 |
| 17p12dup | 9 |
| 17q12del | 4 |
| 17q12dup | 3 |
| 1q21.1del | 6 |
| 1q21.1dup | 3 |
| 22q11.2del | 1 |
| 22q11.2dup | 6 |

(b)

| BLOOD TEST | Estimate | Std Error | t.value | P-value | p.adjust (BH) |
|---|---|---|---|---|---|
| ALBUMIN_BLD | -0.01 | 0.04 | -0.12 | 0.90 | 0.97 |
| ALK_PHOSPHATASE_BLD | 0.02 | 0.05 | 0.42 | 0.67 | 0.91 |
| ALT_SGPT | 0.01 | 0.05 | 0.17 | 0.86 | 0.97 |
| AST_SGOT | -0.03 | 0.05 | -0.57 | 0.57 | 0.91 |
| BASOPHIL | 0.04 | 0.04 | 0.89 | 0.37 | 0.91 |
| BILIRUBIN_TOTAL | 0.01 | 0.04 | 0.27 | 0.78 | 0.91 |
| CALCIUM_BLD | -0.03 | 0.05 | -0.70 | 0.48 | 0.91 |
| CARBON_DIOXIDE_BLD | -0.03 | 0.05 | -0.71 | 0.47 | 0.91 |
| CHLORIDE_BLD | -0.02 | 0.05 | -0.49 | 0.62 | 0.91 |
| CHOLESTEROL | 0.04 | 0.05 | 0.88 | 0.38 | 0.91 |
| CHOLHDL_CHOL_RATIO | 0.09 | 0.05 | 1.78 | 0.07 | 0.75 |
| CREATININE_SERUM | 0.06 | 0.04 | 1.66 | 0.10 | 0.75 |
| EOSINOPHIL | 0.01 | 0.05 | 0.30 | 0.77 | 0.91 |
| GLUCOSE | 0.06 | 0.04 | 1.34 | 0.18 | 0.86 |
| HDL_CHOLESTEROL | -0.05 | 0.05 | -1.09 | 0.28 | 0.87 |
| HEMATOCRIT | -0.03 | 0.04 | -0.71 | 0.48 | 0.91 |
| HEMOGLOBIN | -0.03 | 0.04 | -0.76 | 0.44 | 0.91 |
| LDLHDL_RATIO | 0.07 | 0.05 | 1.44 | 0.15 | 0.82 |
| LDL_CHOLESTEROL | 0.03 | 0.05 | 0.66 | 0.51 | 0.91 |
| LYMPHOCYTE | -0.05 | 0.05 | -1.13 | 0.26 | 0.87 |
| MEAN_CORP_HGB | 0.02 | 0.04 | 0.35 | 0.72 | 0.91 |
| MEAN_CORP_HGB_CONC | -0.01 | 0.04 | -0.36 | 0.72 | 0.91 |
| MEAN_CORP_VOLUME | 0.02 | 0.04 | 0.56 | 0.57 | 0.91 |
| MEAN_PLT_VOLUME | -0.09 | 0.05 | -2.01 | 0.04 | 0.75 |
| MONOCYTE | -0.01 | 0.05 | -0.26 | 0.79 | 0.91 |
| NEUTROPHIL | 0.08 | 0.05 | 1.65 | 0.10 | 0.75 |
| PLATELET | 0.00 | 0.04 | 0.03 | 0.98 | 0.98 |
| POTASSIUMBLD | 0.00 | 0.05 | 0.07 | 0.94 | 0.97 |
| PROTEIN_TOTAL_BLD | 0.00 | 0.05 | -0.07 | 0.95 | 0.97 |
| PROTEIN_TOTAL_SERUM | -0.08 | 0.05 | -1.49 | 0.14 | 0.82 |
| RBC_BLOOD_CELL | -0.05 | 0.04 | -1.11 | 0.27 | 0.87 |
| RED_BLOOD_CELL | -0.02 | 0.04 | -0.55 | 0.58 | 0.91 |
| RED_DISTRIB_WIDTH | 0.04 | 0.04 | 0.88 | 0.38 | 0.91 |
| SODIUM_BLD | -0.03 | 0.05 | -0.68 | 0.49 | 0.91 |
| TRIGLYCERIDES | 0.06 | 0.05 | 1.17 | 0.24 | 0.87 |
| TSH | 0.02 | 0.05 | 0.41 | 0.68 | 0.91 |
| UREA_NITROGEN_BLD | 0.07 | 0.04 | 1.65 | 0.10 | 0.75 |
| WHITE_BLOOD_CELL | -0.01 | 0.05 | -0.31 | 0.76 | 0.91 |
